# White matter tract integrity is reduced in major depression and in individuals with genetic liability for depression

**DOI:** 10.1101/2022.11.09.22282111

**Authors:** David Nothdurfter, Philippe Jawinski, Sebastian Markett

## Abstract

Major depression has been linked to alterations of the brain’s white matter architecture. Major depression has also a considerable genetic component. It is, however, unclear if genetic liability for depression relates directly to the suggested alterations at the brain level.

We re-evaluated white matter fiber tract alterations in major depression and aimed to relate polygenic risk for depression in unaffected individuals to these alterations.

We conducted a population-based retrospective cohort study with N=17183 participants (n=287 currently depressed, n=5536 formerly depressed, n= 13360 healthy) aged 44 to 82 from the UK Biobank imaging cohort, collected between 2015 and 2020.

Depression status was assessed based on questionnaire data. We derived 27 major white matter tracts from diffusion MRI and indexed their integrity according to mean fractional anisotropy. Genetic liability for depression was quantified by polygenic scores based on three recent case-control GWAS.

White matter integrity was globally reduced in depression. The reduction was most pronounced on thalamic and intracortical fiber tracts, where the reduction was observed irrespective of current symptom status. Non-depressed individuals showed already a reduction in relevant fiber tracts with increasing polygenic risk, particularly in thalamic radiations. Subsequent in-silico simulations and non-parametric tests confirmed that the polygenic association was stronger than expected given the low-level statistical dependencies between target and discovery samples. White matter integrity on thalamic and association tracts is reduced in depression irrespective of current disease status and relates to the underlying genetic risk factors in unaffected individuals. The observed statistical links between genes, brain, and disorder adhere to the definition of an intermediate phenotype at the brain level.

## 1. Introduction

Depression is among the most prevalent psychiatric disorders in Europe and one of the leading causes of disability around the globe^1,2^. Recent genome-wide association studies (GWAS) have confirmed that depression is considerably heritable and highly polygenic with many individual genetic variants of very small effect sizes each^3–6^. At the time of writing, about 3.2% of the risk towards major depression can be explained by polygenic scores, i.e., weighted sum scores that aggregate information of multiple trait-associated alleles from GWAS^4^. Little is known, however, about how this genetic risk might translate into depression. In the present study, we aim at identifying brain phenotypes that may mediate the effects of genetic variation on disease risk. Neuroimaging studies suggest alterations of the integrity of white matter pathways in people suffering from depression^7^. While the findings of lower global white matter integrity in people suffering from depression are robust^8^, it remains open which specific brain pathways are altered. Previous research has yielded mixed results, also due to different MRI protocols or limited power in studies with low sample size^9^. Meta-analyses have found most consistently alterations in the genu of the corpus callosum but have produced mixed results regarding other fiber tracts^8–11^. Even larger sample size may be needed to overcome limitations and to resolve inconsistencies regarding implicated fiber tracts. Another reason for inconsistent findings might be heterogeneity in case definition when not distinguishing between currently depressed individuals and individuals with a life-time history of depression. It has been suggested, for instance, that individuals suffering from acute depression exhibit more pronounced alterations of connectivity patterns^9^.

The goals of the present study are twofold. First, we seek to re-evaluate alterations in white matter integrity. By distinguishing between life-time as well as currently depressed individuals in a large imaging sample (UK Biobank with 19,183 participants) we seek to establish both global and local connectivity markers of depression. Second, we seek to link depression-related white matter alterations to the genetic risk for depression in the healthy population. Studies on the relationship between polygenic risk for depression and brain connectivity have yielded divergent results thus far^12,13^ but were also not based on the most recent GWAS findings on depression. Establishing neural markers for depression (goal 1) and linking these markers to polygenic liability (goal 2) are major steps towards a deeper understanding of the etiology of depression^14^.

## 2. Methods

### 2.1. Data resource

All participants were drawn from the UK Biobank (UK Biobank, 2020a). Access to the UK Biobank was granted under project 42032 (PI: Sebastian Markett). We focused on a subset of *N* = 19,183 participants with neuroimaging (T1- and diffusion-weighted) and with complete genotyping data (50.27% men; mean age *M* = 64.08 years (*SD* = 7.52, range: 45 – 82).

### 2.2 Exclusion criteria

We excluded participants with poor quality in the structural T1-weighted image, participants with sex-aneuploidy and whose reported sex did not match their genetic sex, and participants with a history of neurological disorders such as dementia or psychiatric disorders other than depression, social phobia or anxiety, panic attacks, or generalized anxiety disorder. Details on the exclusion procedure are described in the supplementary methods.

### 2.3 Depression status

Depression status was assessed based on participants’ responses to a touchscreen questionnaire presented during the imaging visit^15^. We assigned participants into three groups: *depressed without current symptoms, currently depressed* and *healthy controls*. Details on the assignment to the groups are described in the supplementary methods. In total, depression status was obtained for 19,183 subjects (13360 healthy, 5536 formerly depressed and 287 currently depressed individuals). For descriptive statistics see supplementary table T1.

### 2.4 Group matching

Because measures of white matter integrity are influenced by sex, age and image quality, we performed an additional analysis in a subset of the sample in which the three groups were matched for these variables (via propensity score matching; see supplementary methods). The matched sub-sample included 8653 participants (see figure 1 and supplementary tables T1-T4).

**Figure 1.**
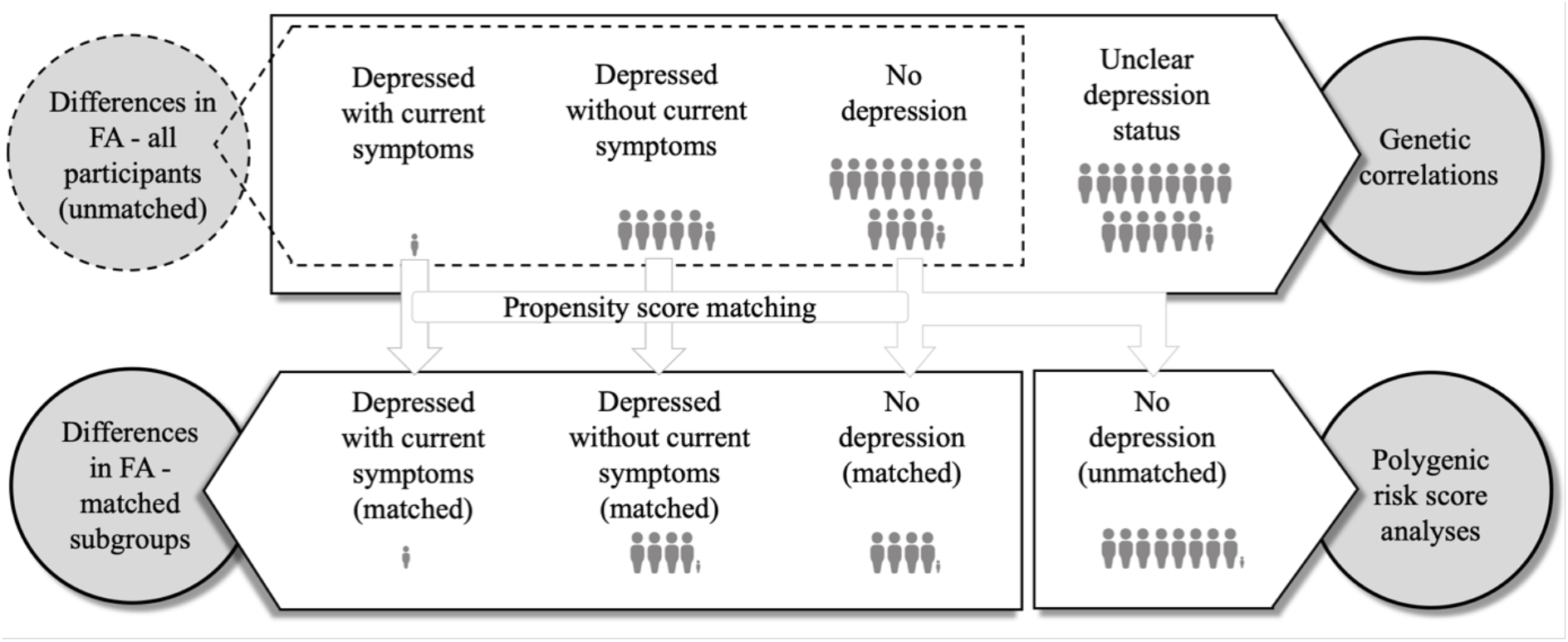
Overview of the sub-samples included for the different analyses. Each depicted person-icon represents 1,000 participants. Smaller icons represent proportionally lower numbers of participants (unmatched sub-samples: depressed with current symptoms 287 participants, depressed without current symptoms 5536 participants, no depression 13360 participants, unclear depression status 15876 participants; matched sub-samples: depressed with current symptoms 286 participants, depressed without current symptoms 4185 participants, no depression 4182 participants; no depression (remaining unmatched) 8092 participants).

### 2.5 MRI analyses

MRI data were acquired on Siemens Skyra 3T scanners at three different imaging facilities across the UK. We used the preprocessed imaging-derived phenotypes for DTI data, generated by an imaging-processing pipeline run by the UK Biobank^16^. In brief, tractography was perfomed in FSL’s probtrackx and 27 major tracts were automatically reconstructed from the data and characterized by mean fractional anisotropy (FA). More details are given in the supplementary methods.

Because fiber tracts share a substantial amount of variance^17^, we computed summary FA measures (via principal component analyses) for groups of fiber tracts (all fiber tracts, association/ commissural fibers, thalamic radiations and projection fibers), following suggestions from previous work^18^ (see supplementary methods for more details).

Multiple regression analyses were then performed to investigate differences in fractional anisotropy between the three groups, individually for the three comparisons (currently vs. formerly depressed, currently depressed vs. healthy, formerly depressed vs. healthy). The four PCs (summary FA measures) were each set as the dependent variable, while depression status served as the independent variable. Also, multiple regressions were performed with FA of individual fiber tracts as dependent variable (applying the Benjamini-Hochberg procedure with a false discovery rate of 5%^19^).

### 2.6. Polygenic risk score analyses

Polygenic risk scores were based on three genome-wide association studies (GWAS)^4–6^. Details on the calculation of the polygenic risk sores are described in the supplementary methods. To investigate if higher polygenic risk for depression is associated with lower white matter integrity on depression-related fiber tracts, we analyzed the remaining healthy participants that were not used for the previous analyses with the matched subgroups. To ensure full equivalence between this analysis and the group comparison, we used PCs from the group comparison and projected the unmatched healthy individuals’ data onto these components. The resulting factor scores as well as FA values of individual fiber tracts were then correlated with participants’ polygenic risk scores for depression. Multiple regressions were performed with factor scores as dependent variable and each polygenic risk score as independent variables. In addition to the above-mentioned covariates, genotyping array and 20 genetic principal components were included. As the three GWAS discovery samples showed varying sample overlap (6.19, 44.74 and 59.58%) with the full UKB cohort from which we selected a subsample for the present study, we performed additional analyses to ensure that the overlap does not confound our conclusions. We benchmarked our results in comparison to a null model that was based on a series of simulations in which we explored how random variables with varying intercorrelations with depression status are related to the polygenic risk scores. The rationale behind these analyses was to quantify the impact of sample overlap between GWAS discovery and target samples under different boundary conditions. As part of this benchmarking, each empirical PGS association was compared to the distribution of PGS associations with 10,000 random synthetic variables which were similarly intercorrelated with depression status as white matter integrity. We refer to the supplementary material for more details.

### 2.7. Genetic correlations

We conducted GWAS using *PLINK2* for the FA phenotypes that differed between groups. All participants with available genetic and imaging data were included for this step. We then performed linkage disequilibrium score regression to calculate SNP-based heritabilities^20^. Finally, genetic correlations were performed between the depression phenotypes and the FA phenotypes.

### 2.8. Covariates and Outlier exclusion

Highest degree of education, intake of antidepressants, sex, age, age^2^, image quality and the location of the assessment center were entered as covariates (see supplementary methods for more details).For all analyses, we excluded participants with outliers in the PC or FA value of interest by applying the Tukey criterion. On average, 1.27% (*SD=*0.76) of the participants were excluded in the different analyses due to outliers. Figure 1 provides an overview of the different sub-samples of the UK Biobank used for the various analyses.

### 2.9 Data and Code availability statement

All analyses were performed in *R* version 3.6.3. All data were taken from the UK Biobank. Data access is limited to registered researchers via https://www.ukbiobank.ac.uk/. We will publish analysis code for the null model for PGS testing as well as code to reproduce the figures on the open science framework upon final acceptance of the manuscript.

## 3. Results

### 3.1. Differences in global FA components

We first investigated group differences in global FA as quantified by the first principal component (PC) across all fiber tracts and across all fiber tract groups. Supplementary tables T5 and T6 depict loadings of individual fiber tracts on these PCs. Figure 2 shows the comparison of the four fiber groups’ PCs for the subgroup of matched participants. Multiple regression analyses revealed that depressed participants without current symptoms showed significantly lower values on the PC derived from all fiber tracts (*b* = −0.184, *p* = 0.008), association/commissural fibers and thalamic radiations than healthy participants (*b* = −0.219, *p* = .018; *b* = −0.132, *p* < .001). Between healthy participants and depressed participants with current symptoms, the PCs derived from association/commissural fibers and thalamic radiations differed significantly (*b* = −0.38, *p* = .019; *b*= −0.252, *p* = .023). No differences in any of the four PCs could be observed between the two depression groups. The results were similar to the analyses including the full sample, except for a lack of significant difference in thalamic radiations between currently depressed and healthy participants (see tables T7 to T9).

**Figure 2.**
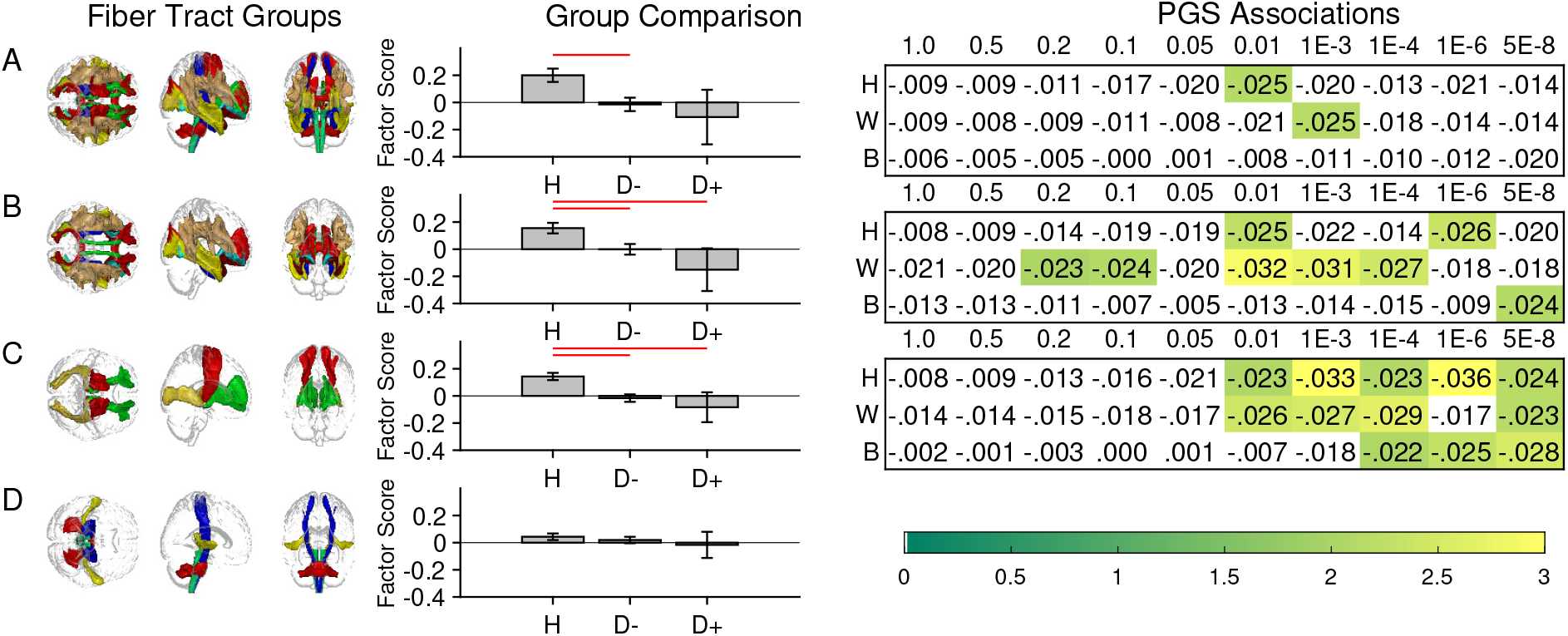
Overall integrity of all white matter fiber tracts (A) was reduced in individuals with a history of depression but no current symptoms (D-) relative to healthy participants (H). For association fiber tracts (B) and thalamic radiations (C), the reduction was additionally visible for the smaller group of individuals with current symptoms (D+). No such difference was observed for projection fibers (D). The bar graphs in the middle column give the factor scores of the fiber tracts in the matched subgroups (caliper 0.05). The red lines mark significant group differences. The heatmaps on the right list association strengths (standardized beta coefficients) between fiber tract integrity and polygenic scores based on the GWAS by Howard et al. (H), Wray et al. (W), and Baselmans et al. (B) across different thresholds. Significant coefficients are highlighted according to the corresponding t-statistic.

### 3.2. Differences in individual fiber tracts

Eleven individual fiber tracts showed decreased FA in depressed participants without current symptoms in at least the full sample or the sub-sample including matched participants, compared to healthy participants (figure 2). Between healthy participants and depressed participants with current symptoms significant differences in FA were found for five fiber tracts (figure 2). None of the fiber tracts differed between the two depression groups.

### 3.3. Polygenic risk scores

Figure 2 depicts the associations between PGS and fiber integrity in healthy participants of the fiber tract groups that were linked to depression in the group comparison (all fiber tracts, commissural/association fibers, thalamic radiations). With increasing genetic liability for depression (higher PGS scores), healthy individuals showed reduced integrity of commissural/association fiber tracts and of the thalamic radiations, suggesting a subclinical alteration of white matter integrity. Of note, this relationship was observed for all three operationalizations of polygenic risk and extended across several PGS thresholds. Figure 3 depicts the association of PGS and the integrity of the individual fiber tracts for which we observed group differences between healthy participants and one of the two depression groups. Every fiber tract except the left inferior longitudinal fasciculus showed significant association with at least one of the three PGS, most consistently for the anterior thalamic radiations and the forceps minor.

**Figure 3.**
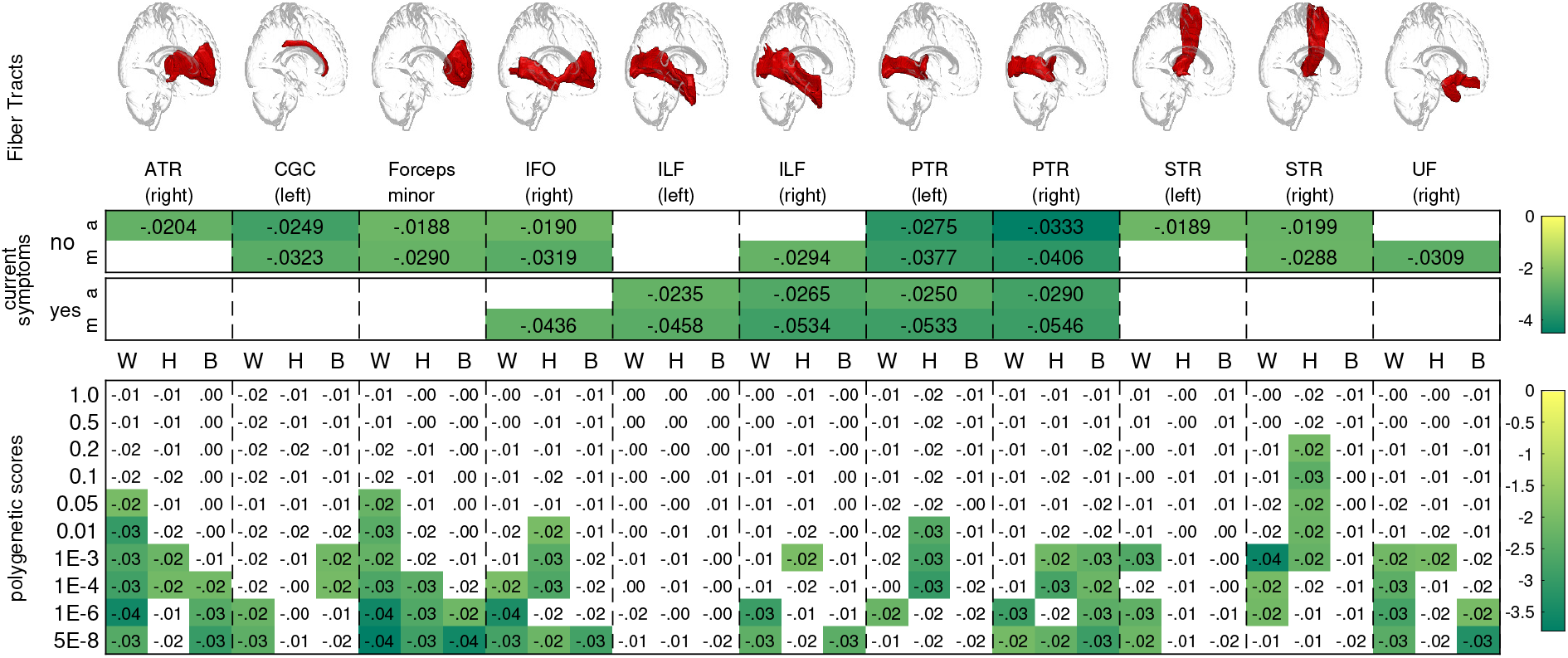
Eleven fiber tracts (top panel) showed group differences individually (middle panel). We present standardized beta coefficients for group differences between healthy individuals and individuals with a history of depression (no current symptoms) and individuals with current symptoms, based on matched groups (m) and all participants (a). Only FDR-significant coefficients are given and color-coded by the corresponding t-value. Polygenic score associations are depicted in the bottom panel (different discovery GWAS in columns, different thresholds in rows). Only significant associations are highlighted and color coded by the corresponding t-value.

### 3.4 Genetic correlations

We sought to corroborate previous heritability estimates of depression through LD score regression: Depending on the summary statistics from the original GWAS, heritability estimates were 9.6%^6^, 6.3%^4^, and 3.1%^5^. SNP-based heritability for the global FA components was considerable, ranging between 29-30% (supplementary table T21). Genetic correlations between depression and global FA components were all in the expected directions but not statistically significant (supplementary table T21). SNP-based heritability for individual fiber tracts ranged between 19 and 33%. Genetic correlations with depression were all in the expected direction but did not survive the correction for multiple comparisons (see supplementary table T22).

### 3.5 Validation analysis

To investigate the influence of partly overlapping participants between the present and the GWAS discovery samples, we simulated synthetic random data for which we systematically manipulated the degree of intercorrelation with actual depression status. We then ran partial correlation analyses identical to our actual analysis of white matter fiber statistics (see methods and supplementary methods). As expected, we observed a spurious increase in polygenic association with increasing intercorrelations between depression status and random data when including patients and controls (Figure 4A). When limiting the analysis to healthy controls, however, the partial correlations where close to zero (< |1e-15|), irrespective of the degree of intercorrelation between the synthetic data and depression status (figure 4B). While the simulation results suggest that partially overlapping discovery and target samples do not affect the validity of PGS associations in a scenario where GWAS summary statistics are obtained by case-control comparisons and the target sample includes only controls, we still decided to complement the simulations with a further validation step: For each fiber group, we established a null model of 10,000 random variables whose intercorrelation with depression status matched the empirical correlations between fiber integrity and depression status. If the here reported PGS associations with fiber integrity were simply a result of the partially overlapping samples, we would except similar PGS associations with the random data. This, however, was not the case: For association/commissural tracts and for the thalamic radiations, we confirmed significant reductions in fiber integrity in healthy participants with higher genetic liability for depression (figure 4C). Only for the global measure of white matter integrity of all fiber tracts, the comparison to the null model did not confirm the descriptive PGS association, suggesting that the observed correlation is not stronger than the inherent interdependencies resulting from sample overlap.

**Figure 4.**
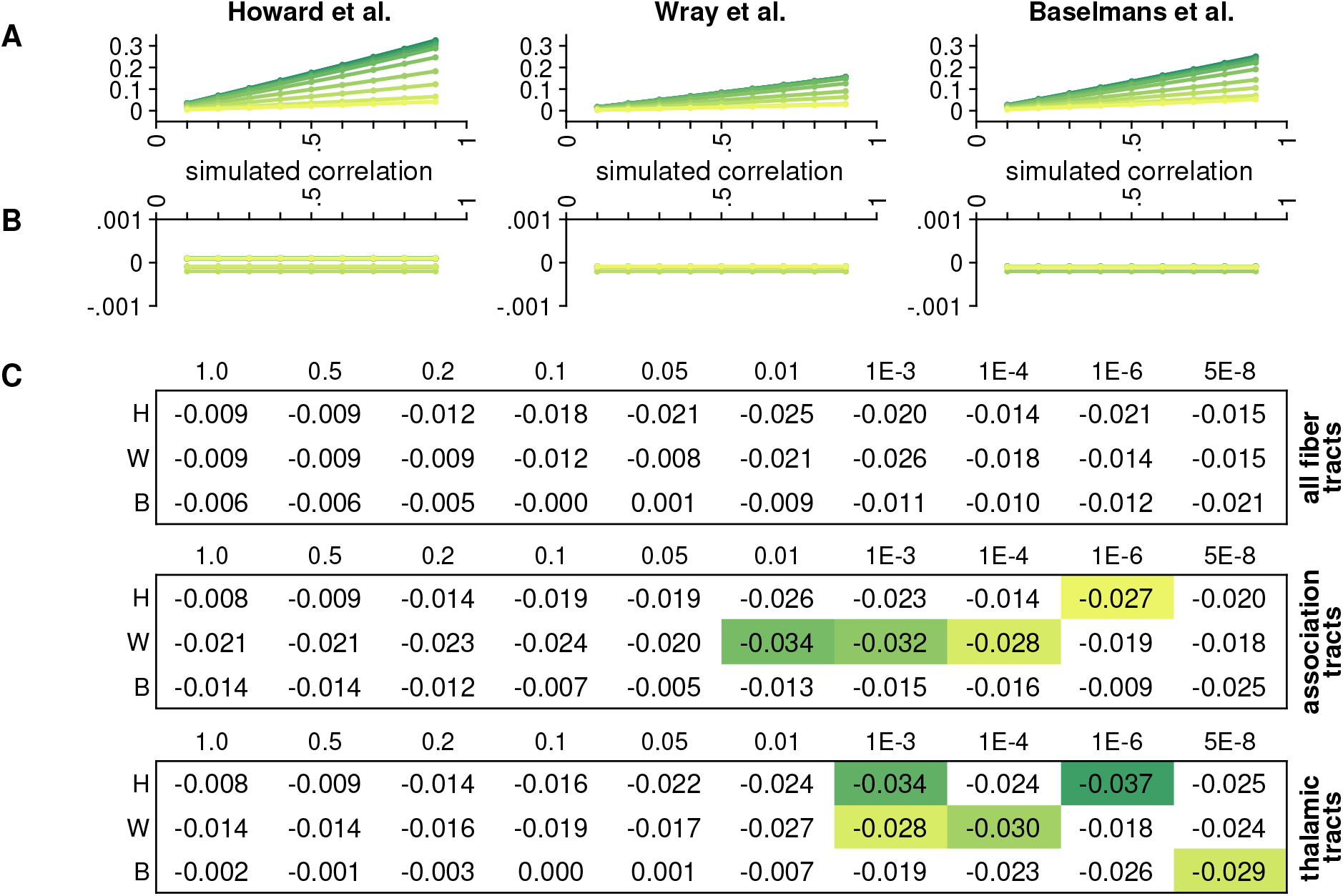
Results from the validation analyses: PGS associations with synthetic data are only substantial when the target sample includes cases and controls, and increase as a function of similarity between the variable of interest and disease status (A). When only healthy participants are included in the target sample, the PGS associations with random data is practically zero (B). The different lines reflect PGS on different thresholds, with more yellow hues reflecting less liberal thresholds. The highlighted empirical PGS associations in panel C are significantly larger (i.e. more negative) than PGS associations with comparable random data (p<.05, non-parametric test with 10,000 random variables).

## 4. Discussion

The present study aimed at establishing structural connectivity markers for major depression, and linking depression-related white matter alterations to the genetic risk for depression. We found lower white matter fiber tract integrity in depression using data from the UK Biobank, extending previous work with an earlier data release^18^. Our results support previous findings on reduced integrity of interhemispheric fiber tracts, particularly the forceps minor^8–11,21^, fiber tracts belonging to the Papez circuit which plays a role in negative affect^22^, and the medial forebrain bundle which includes the cingulum bundle and the uncinate fasiculus^23^ (Bracht et al., 2015). We further observed that similar reductions in white matter integrity present already subclinically in healthy individuals with higher genetic liability for depression.

Mental disorders are considerably heritable^24^ and contemporary GWAS results can be used to quantify an individual’s genetic liability for a disorder in the form of polygenic scores^25^. Linking PGS to brain structure and function bears the potential to elucidate the neural mechanisms by which genetic risk translates into the disorder, to assess interactions with environmental factors, and to characterize sub-clinical expression in unaffected populations. So far, PGS-brain relationships have mostly been investigated in the context of schizophrenia and autism^1^ but not all attempts have been successful^31,32^. PGS for depression and related phenotypes have also been linked to brain structure and function^33–36^, but not all studies have distinguished between cases and controls, or have linked the PGS-brain associations to case/control differences. Particularly the latter, however, is crucial to establish links between genes, brain, and behavior, and to establish intermediate phenotypes for the disorder. To the best of our knowledge, our current results are the first to confirm neurostructural differences in individuals with depression and link the same neurostructural variables to genetic liability for depression in unaffected individuals. Our results thus provide major implications for structural connectivity as an intermediate phenotype for depression.

To qualify as an intermediate phenotype (or endophenotype) of depression, an imaging-derived phenotype needs to be related to the disorder, needs to be heritable, and needs to be relatable to the genetic underpinnings of the disorder in unaffected individuals^14^. We confirmed these criteria for white matter integrity: Table 1 summarizes our findings regarding these criteria for different groups of fiber tracts. Using LD-score regression, we confirmed a SNP-based heritability for white matter tract integrity ranging from 19 and 33%. Using polygenic scores, we confirmed that genetic liability for depression is not only associated with the disorder itself, but also with white matter tract integrity, which is a key postulate for an endophenotype^37,38^. Even though total effect sizes were low, the significant reductions in white matter integrity with increasing genetic risk load in healthy individuals indicate that white matter integrity of implicated fiber tracts fulfill the endophenotype criteria. The small effect sizes are a likely consequence of the high polygenicity of the genetic underpinnings of depression and the limited prediction accuracy of polygenic scores to date.^39^

**Table 1.**
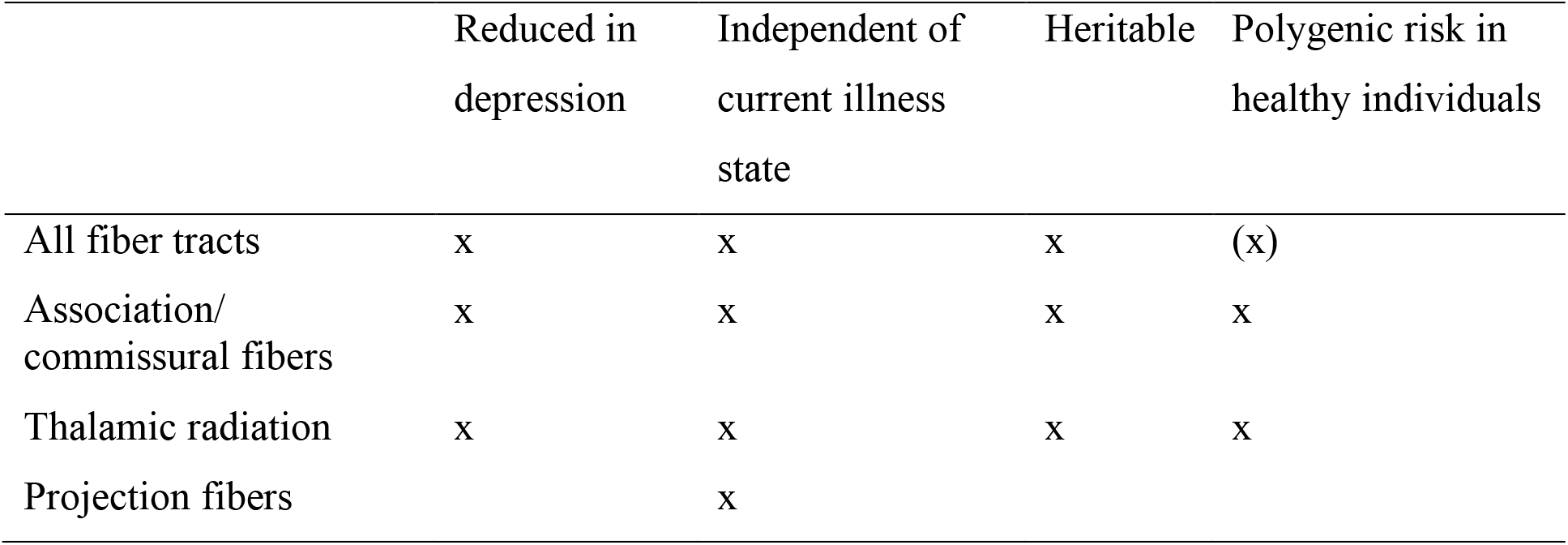
Summary of the main results with respect to the endophenotype concept. White matter integrity of association and commissural fiber tracts is heritable, reduced in depression independent of current symptoms, and subclinically reduced in healthy individuals who are genetically at risk.

|  | Reduced in<br>depression | Independent of<br>current illness<br>state | Heritable | Polygenic risk in<br>healthy individuals |
| --- | --- | --- | --- | --- |
| All fiber tracts | x | x | x | (x) |
| Association/<br>commissural fibers | x | x | x | x |
| Thalamic radiation | x | x | x | x |
| Projection fibers |  | x |  |  |

A well-known pitfall in PGS analyses is the spurious increase in association strength when the GWAS discovery sample and the target sample overlap^40^. Even partial sample overlap can dramatically inflate observed associations. Partially overlapping samples, however, should be less of a concern when the GWAS summary statistics reflect a group comparison (such as individuals with depression vs. healthy controls) and the target variable of interest is only assessed in individuals belonging to either group (such as healthy participants only). We corroborated this conjecture through a series of simulations. And we established through a simple non-parametric test that the observed relationship between polygenic liability for depression and white matter integrity of intracortical and thalamic fiber tracts is substantial, even in the face of overlapping samples. Given the large sample size requirements of discovery GWAS^39^, sample overlap will remain a concern, at least until the predictive accuracy of PGS will improve to the point that meaningful associations can already be observed in small target samples^41^. The here presented non-parametric test does not depend on prior knowledge of sample overlap, does not require access to individual-level data from the discovery cohort, and is easy to implement, particularly in psychiatric research contexts where summary statistics often result from case/control comparisons.

Contrary to the polygenic score analyses, genetic correlations did not reveal significant associations between FA phenotypes and depression. This may be explained by the lower statistical power of summary-based (genetic correlations via LD score regression) relative to individual-level association analysis (polygenic score analysis). Findings from genetic correlations may indicate that the total genetic overlap between FA phenotypes and depression is modest, in line with previous findings linking psychiatric diseases to brain structure^42^. The lack of significant genetic correlations between FA and depression needs further investigation. Another general limitation to the present study is the group definition which was not based on structured clinical interviews but on self-report data. Even though this assessment led to similar prevalence rates of depression as other studies^15^, it is still possible that depression in the present study may have been over-diagnosed.

In summary, we report structural connectivity differences between healthy and depressed individuals. These alterations are present in currently depressed as well as formerly depressed individuals, are heritable, and seem to be related to higher genetic liability for depression in healthy individuals. Our results thus provide indications for an involvement of structural connectivity in the genetic etiology of depression. Identifying molecular substrates of brain connectivity may thus reflect a promising strategy to inform individualized treatment^43–45^ and improved diagnostics in the near future^46,47^

## Data Availability

All data produced are available online at ukbiobank.ac.uk.

