## Supplementary Materials for "White matter tract integrity is reduced in major depression and in individuals with genetic liability for depression"

**David Nothdurfter, Philippe Jawinski, Sebastian Markett**
**Humboldt-Universität zu Berlin**

### 1. Supplementary Methods

#### *1.1. Exclusion criteria and covariates*

Imaging quality was assessed by noise contrast rating, inhomogeneity contrast ratio, and resolution rating (per voxel fraction root mean square (RMS) error) as implemented in the Computational Anatomy Toolbox CAT12 (http://www.neuro.uni-jena.de/cat/). A quality rating of > 3 was set as threshold. If participants were related to each other (kinship coefficient < 0.0443), only one of them was included in the analysis.

If participants' hospital inpatient data (datafield 41270), data about self-reported diseases (datafield 20002) or their mental health questionnaire (datafield 20544) included any psychiatric disorders other than depression, social phobia or anxiety, panic attacks and generalized anxiety disorder, they were excluded from the analysis.

Participants answered a set of questions to screen for bipolar disorder during the imaging visit (see Smith et al., 2016). If they fulfilled the criteria for a bipolar disorder as described in Smith et al. (2016), they were excluded from this study.

In addition to psychiatric disorders, participants with multiple sclerosis, Parkinson's disease, dementia, Alzheimer's, cognitive impairment, other demyelinating disease (not multiple sclerosis), neurological injury/trauma, chronic/degenerative neurological problem or head injury were excluded from the analysis. These diagnoses were assessed via hospital inpatient data and self-reported diseases (datafields 41270 and 20002). Participants with the following ICD-10 diagnoses were excluded from the analyses:

F00, F000, F001, F002, F009, F01, F010, F011, F012, F013, F018, F019, F02, F020, F021, F022, F023, F024, F028, F03, F06, F060, F061, F062, F063, F064, F065, F066, F067, F068, F069, F070, F078, F079, F09, F102, F112, F122, F132, F142, F162, F182, F192, F20, F200, F201, F202, F203, F204, F205, F206, F208, F209, F21, F22, F220, F228, F229, F23, F230, F231, F232, F233, F238, F239, F25, F250, F251, F252, F258, F259, F28, F29, F30, F301, F302,

F308, F309, F31, F310, F311, F312, F313, F314, F315, F316, F317, F318, F319, F44, F441, F442, F443, F444, F445, F446, F447, F448, F449, F840, F841, F45, F450, F451, F452, F453, F453, F454, F458, F459, F48, F480, F481, F488, F489, F60, F601, F602, F603, F604, F605, F606, F607, F608, F609, F61, F70, F701, F702, F708, F709, F71, F710, F711, F718, F719, F72, F720, F721, F728, F729, F73, F730, F731, F738, F739, F78, F780, F781, F788, F789, F79, F790, F791, F798, F799.

Additionally, for the never-depressed group participants with the following diagnoses were also excluded: F32, F320, F321, F322, F323, F328, F329, F33, F331, F332, F333, F334, F338, F339, F34, F340, F341, F348, F349, F38, F380, F381, F388, F39, F40, F400, F401, F403, F408, F409, F41, F410, F411, F412, F413, F418, F419, F42, F420, F421, F422, F428, F429, F431.

Antidepressants intake was confirmed if participants took any of the following medications (all antidepressants currently available in the United Kingdom): agomelatine, amitriptyline, citalopram, clomipramine, desipramine, dosulepin, doxepin, duloxetine, escitalopram, fluoxetine, fluvoxamine, imipramine, isocarboxazid, lofepramine, mianserin, mirtazapine, moclobemide, nortriptyline, paroxetine, phenelzine, reboxetine, sertraline, tranylcypromine, trazodone, trimipramine, venlafaxine, vortioxetine.

#### *1.2. Group assignment*

Participants in the group *depressed without current symptoms* had experienced feelings of depression or anhedonia for at least two weeks once in their life, had seen a general practitioner or psychiatrist for ‘nerves, anxiety or depression’, and had experienced depressive symptoms only “several days” or “not at all” in the 14 days prior to the imaging visit. Participants in the *currently depressed group* had felt depressed or experienced anhedonia “nearly every day” in the last 14 days and additionally affirmed to having seen a general practitioner or psychiatrist for nerves, anxiety or depression. Participants were classified in the *healthy control group* if they did not fulfill all of the above described criteria for a history of depression and if they only felt down for “several days” or “not at all” in the last 14 days prior to the imaging visit. Participants were excluded from the healthy control group if they reported ever getting any psychiatric diagnosis by a professional, if their hospital inpatient data included any psychiatric disorder, or if depression was part of their self-reported diagnoses.

#### *1.3. MRI processing and calculation of PCs*

Diffusion data (2 mm isotropic voxels, 104*104*72 matrix, TR = 3600 ms, TE = 92 ms, multiband factor 3) were acquired with diffusion gradients in 100 distinct directions (50 with b = 1000 s/m$^2$ and 50 with b = 2000 s/m$^2$) in anterior-posterior direction.

DTI data were first corrected for eddy currents and head motion by use of FSL's Eddy Tool. Then, tractography-based analysis was performed by first modelling within-voxel multi-fiber tract orientation structure using the BEDPOSTx tool. Probabilistic tractography was performed via PROBTRACKx (Behrens et al., 2007; Jbabdi et al., 2012). The pipeline mapped 27 major fiber tracts using standard-space start/stop ROI masks generated by AutoPtx (de Groot et al., 2013). The weighted-mean value of fractional anisotropy within each of these tracts was used as an indicator for structural connectivity in this study - the weighting was determined by the tractography output (see Smith, Almagro, & Miller, 2020).

A global FA component was extracted from the 27 tracts of interest via principal component analysis (PCA) and factor scores of the first principal component (PC) were used to quantify global fiber integrity. The same procedure was implemented for association/ commissural fibers, thalamic radiations and projection fibers, respectively. If fiber tracts loaded negatively on the first component of the PCAs, PCs were sign-adjusted, so that higher factor scores represent higher fiber tract integrity.

#### *1.4. Matching procedure*

Matching was conducted via propensity score matching using the *R* package *MatchIt.* To keep a high percentage of data, groups were not matched 1:1, as each included a different number of participants (287, 5536 and 13360 for the depressed group with and without current symptoms and the healthy group, respectively). The currently depressed group was matched 1:16 with the depressed group without current symptoms and the resulting matched group (depressed participants without current symptoms) was matched 1:1 with the healthy control group. *Nearest neighbor* in R packages *MatchIt* was used as matching algorithm. *Nearest neighbor* matching selects the best control(s) for each treatment unit, i.e. the match with the closest distance measure (logit). For this method, different calipers can be specified – when setting a caliper, only those control units are selected that are within a certain number of standard deviations of the distance measure (the caliper) from the to be matched unit (Ho et al., 2013). The matching was performed using 46 different calipers (between 0.05 and 0.50 with an interval of 0.01). The matching algorithm yielding the highest percent balance improvement (nearest neighbor with caliper 0.05) was selected for the subsequent analyses. However, the global PC

extracted from all fiber tracts was also analyzed with the other matching outcomes to ensure robustness of the results.
For the outcome of every caliper, a multiple regression with the PC extracted from all fiber tracts as dependent variable was performed. Of these 46 regressions, 26 (56.52 %) revealed significantly lower values on the PC extracted from all fiber tracts in depressed participants without current symptoms than in healthy participants ($p < .05$). For 9 other regressions, the *p* value was below .10.

For the comparison of healthy and depressed individuals with current symptoms, two of the regressions yielded a significant difference, for 20 regression the *p* values was below .10. None of the comparisons between depressed participants with and without current symptoms showed a significant difference. For a detailed overview of the various outcomes see table T6.

#### *1.5. Calculations of polygenic risk scores*

*PrSice 2* (Choi et al., 2019) was used to calculate polygenic risk scores for the UK Biobank sample. If base files included info scores or minor allele frequencies (MAF), only variants with an info score >= 0.9 and a MAF >= 0.05 were included. Also, variants in linkage disequilibrium were clumped, so that only independent variants remained – the variant with the lowest *p* value in the base file was selected. Polygenic risk scores were calculated for ten different *p* value thresholds (5e-08, 1e-06, 0.0001, 0.001, 0.01, 0.05, 0.1, 0.2, 0.5, 1) and then *z* standardized. Polygenic risk scores based on Baselmans et al. (2019) were inverted to match the other two polygenic risk scores, because the originally reported summary statistics in the paper are poled towards well-being instead of depression.

#### *1.6. Analyses accounting for sample overlap*

The three GWAS discovery samples showed varying sample overlap (6.19, 44.74 and 59.58%) with the full UKB cohort from which we selected a subsample for the present study. Even partial overlap between the GWAS discovery and the PGS target sample can dramatically inflate associations (Wray et al., 2015). This, however, should only apply for cases where the same phenotype is studied in the discovery and the target sample, and where the GWAS summary statistics are derived from a case/control comparison, and where both cases and controls are included in the target sample. Since our present design focused on a different phenotype (white matter integrity instead of depression status) in healthy individuals, the partial sample overlap should be less of a concern. To affirm this conjecture we ran a set of simulations that explored the relationship between polygenic scores and synthetic variables with varying intercorrelations with depression status in a sample that contained either depression cases and controls or controls only. Using the *R* package *faux*, we created normally-distributed random variables that correlated with depression status (healthy vs. life-time depressed; *r* between 0.1 and 0.9 in an interval of 0.1). We created 10,000 different random variables for each correlation strength. We followed a 10-fold cross-validation scheme and calculated partial correlations (controlling for the same variables as in the main analysis) between polygenic risk scores and theses variables in a random draw of 10 % of the sample (both depressed without current symptoms and healthy participants) as well as in a healthy subsample.

We observed a spurious increase in polygenic associations when including both patients and controls. With increasing intercorrelations between depression status and random data, partial correlations between polygenic risk scores and simulated random variables increased as well. This was particularly pronounced for the polygenic scores derived from GWAS with higher sample overlap (for the different polygenic risk thresholds: *r* between 0.032 and 0.173 for Wray et al. (2018), *r* between 0.046 and 0.362 for Howard et al. (2019), and *r* between 0.059 and 0.279 for Baselmans et al. (2019)). However, such artificial inflation was not apparent when we limited our analysis to healthy controls. In this case, all partial correlations were around zero irrespective of the correlation strength between depression status and the random data (all *r'*s < |1e-15|; see tables T15 to T20 and figure S2).

In a second validation step, we created 10,000 random variables that correlated to the same extent with depression status as the empirical partial correlations found between polygenic risk and white matter integrity (for all tract groups that differed between groups). We then calculated the partial correlations between these simulated variables and polygenic risk for depression in the healthy group. This resulted in 10,000 partial correlations (reflecting PGS association with random data) for each fiber tract group and for each of the three PGS (based on three discovery GWAS). We then determined in how many cases the empirical PGS association with white matter integrity was more substantial (i.e. more negative) than the random association, and calculated a p-value, reflecting the probability to observe a similar correlation in meaningless (i.e. random) data with a similar dependency structure. Results are presented in main figure 4C and in supplementary figure S3.

**Table T1** Distribution of age, sex, education level, image quality rating, intake of antidepressants, assessment center and FA components for all participants

| | **Healthy participants** (*N* = 13360) | | | | **Depressed participants without current symptoms** (*N* = 5536) | | | | **Depressed participants with current symptoms** (*N* = 287) | | | |
|---|---|---|---|---|---|---|---|---|---|---|---|---|
| | *M*/Percent | *SD* | Median | Range | *M*/Percent | *SD* | Median | Range | *M*/Percent | *SD* | Median | Range |
| Sex: Men | 55.60 % | | | 0/1 | 38.30 % | | | 0/1 | 33.40 % | | | 0/1 |
| Age (in years) | 64.81 | 7.52 | 65.53 | 46-82 | 62.51 | 7.23 | 62.42 | 45-81 | 60.65 | 7 | 59.80 | 48-80 |
| Education level | | | | | | | | | | | | |
| Incomplete | 6.3 % | | | | 4 % | | | | 8.7 % | | | |
| Compulsory | 12.9 % | | | | 11.6 % | | | | 13.6 % | | | |
| Continued | 6.1 % | | | | 6.1 % | | | | 5.2 % | | | |
| College | 26.1 % | | | | 24.4 % | | | | 28.2 % | | | |
| University | 48.6 % | | | | 53.9 % | | | | 44.3 % | | | |
| Image Quality Rating | 2.05 | 0.16 | 2 | 1.67-2.99 | 2.03 | 0.14 | 1.99 | 1.62-2.99 | 2.04 | 0.17 | 1.99 | 1.86-2.87 |
| Antidepressants intake | 0.9 % | | | | 12.2 % | | | | 25.8 % | | | |
| Assessment center | | | | | | | | | | | | |
| Cheadle | 60.59 % | | | | 62.57 % | | | | 59.23 % | | | |
| Reading | 13.93 % | | | | 12.05 % | | | | 14.29 % | | | |
| Newcastle | 25.48 % | | | | 25.38 % | | | | 26.48 % | | | |
| Principal component (all fiber tracts) | -0.12 | 3.12 | -0.2 | -8.76 - 8.53 | -0.09 | 3.07 | -0.19 | -8.43-8.29 | -0.09 | 3.22 | -0.49 | -8.42-8.77 |
| Principal component (association/ commissural) | -0.06 | 2.5 | -0.14 | -6.95-6.92 | -0.12 | 2.46 | -0.17 | -6.81-6.71 | -0.09 | 2.53 | -0.49 | -6.95 – 7.13 |
| Principal component (thalamic) | -0.09 | 1.71 | -0.13 | -4.81-4.67 | -0.02 | 1.7 | -0.05 | -4.66-4.64 | -0.01 | 1.73 | 0.02 | 4.43-4.56 |
| Principal component (projection) | -0.08 | 1.51 | -0.1 | -4.21-4.05 | 0.06 | 1.46 | 0.05 | -3.93-4.06 | 0.12 | 1.54 | 0.22 | -3.65-4.26 |

**Table T2** Results of *nearest neighbor* matching with different calipers

| Caliper | percent balance improvement 1 | percent balance improvement 2 | $b_1$ | $p_1$ | $b_2$ | $p_2$ | $b_3$ | $p_3$ |
|---|---|---|---|---|---|---|---|---|
| 0.05 | 99.175 | 99.057 | -0.184 | 0.008 | -0.372 | 0.067 | -0.114 | 0.545 |
| 0.06 | 99.124 | 98.702 | -0.103 | 0.129 | -0.350 | 0.078 | -0.138 | 0.464 |
| 0.07 | 98.738 | 98.276 | -0.107 | 0.112 | -0.208 | 0.294 | -0.103 | 0.581 |
| 0.08 | 98.634 | 98.119 | -0.256 | 0.000 | -0.431 | 0.032 | -0.101 | 0.592 |
| 0.09 | 98.588 | 98.052 | -0.108 | 0.096 | -0.349 | 0.072 | -0.166 | 0.356 |
| 0.10 | 98.253 | 97.781 | -0.235 | 0.000 | -0.352 | 0.069 | -0.097 | 0.593 |
| 0.11 | 97.682 | 97.551 | -0.110 | 0.095 | -0.227 | 0.248 | -0.125 | 0.496 |
| 0.12 | 97.504 | 97.038 | -0.095 | 0.151 | -0.241 | 0.218 | -0.166 | 0.365 |
| 0.13 | 97.035 | 96.665 | -0.162 | 0.011 | -0.360 | 0.059 | -0.138 | 0.440 |
| 0.14 | 96.966 | 96.506 | -0.021 | 0.749 | -0.166 | 0.393 | -0.132 | 0.463 |
| 0.15 | 97.097 | 96.187 | -0.107 | 0.099 | -0.248 | 0.202 | -0.124 | 0.492 |
| 0.16 | 95.828 | 95.801 | -0.186 | 0.004 | -0.351 | 0.066 | -0.122 | 0.496 |
| 0.17 | 95.073 | 95.524 | -0.181 | 0.005 | -0.320 | 0.101 | -0.114 | 0.532 |
| 0.18 | 96.003 | 95.517 | -0.159 | 0.015 | -0.281 | 0.142 | -0.120 | 0.518 |
| 0.19 | 95.986 | 94.862 | -0.162 | 0.011 | -0.286 | 0.135 | -0.105 | 0.556 |
| 0.20 | 95.092 | 94.692 | -0.131 | 0.046 | -0.334 | 0.091 | -0.126 | 0.487 |
| 0.21 | 93.491 | 94.226 | -0.173 | 0.007 | -0.371 | 0.052 | -0.151 | 0.405 |
| 0.22 | 92.860 | 93.884 | -0.147 | 0.024 | -0.343 | 0.077 | -0.140 | 0.442 |
| 0.23 | 93.256 | 93.778 | -0.073 | 0.265 | -0.248 | 0.209 | -0.157 | 0.390 |
| 0.24 | 92.309 | 93.345 | -0.063 | 0.330 | -0.255 | 0.187 | -0.177 | 0.339 |
| 0.25 | 91.796 | 93.065 | -0.163 | 0.014 | -0.402 | 0.042 | -0.165 | 0.377 |
| 0.26 | 90.984 | 92.403 | -0.108 | 0.093 | -0.325 | 0.095 | -0.138 | 0.447 |
| 0.27 | 90.966 | 91.814 | -0.132 | 0.039 | -0.228 | 0.233 | -0.108 | 0.548 |
| 0.28 | 91.122 | 91.910 | -0.100 | 0.121 | -0.307 | 0.112 | -0.140 | 0.436 |
| 0.29 | 90.341 | 91.143 | -0.129 | 0.051 | -0.334 | 0.092 | -0.151 | 0.417 |
| 0.30 | 88.890 | 90.947 | -0.078 | 0.233 | -0.243 | 0.216 | -0.138 | 0.453 |
| 0.31 | 87.539 | 90.736 | -0.098 | 0.125 | -0.308 | 0.109 | -0.141 | 0.437 |
| 0.32 | 86.975 | 91.442 | -0.213 | 0.001 | -0.365 | 0.061 | -0.102 | 0.574 |
| 0.33 | 85.661 | 89.960 | -0.117 | 0.062 | -0.214 | 0.260 | -0.098 | 0.579 |
| 0.34 | 87.167 | 89.892 | -0.157 | 0.017 | -0.302 | 0.125 | -0.119 | 0.517 |
| 0.35 | 85.833 | 89.756 | -0.123 | 0.060 | -0.298 | 0.130 | -0.112 | 0.548 |
| 0.36 | 85.063 | 88.465 | -0.105 | 0.110 | -0.271 | 0.175 | -0.139 | 0.448 |
| 0.37 | 85.365 | 87.365 | -0.128 | 0.048 | -0.344 | 0.078 | -0.167 | 0.361 |
| 0.38 | 84.088 | 87.085 | -0.117 | 0.066 | -0.335 | 0.078 | -0.129 | 0.474 |
| 0.39 | 82.910 | 88.565 | -0.131 | 0.043 | -0.322 | 0.099 | -0.129 | 0.478 |
| 0.40 | 82.484 | 87.119 | -0.156 | 0.015 | -0.314 | 0.109 | -0.136 | 0.448 |
| 0.41 | 82.294 | 86.514 | -0.177 | 0.007 | -0.359 | 0.067 | -0.130 | 0.493 |
| 0.42 | 81.542 | 86.643 | -0.139 | 0.032 | -0.268 | 0.172 | -0.116 | 0.525 |
| 0.43 | 81.326 | 86.562 | 0.174 | 0.007 | 0.324 | 0.096 | 0.082 | 0.652 |
| 0.44 | 81.600 | 86.074 | 0.143 | 0.027 | 0.283 | 0.147 | 0.167 | 0.360 |
| 0.45 | 78.784 | 85.010 | -0.164 | 0.012 | -0.271 | 0.169 | -0.128 | 0.490 |
| 0.46 | 79.407 | 83.940 | -0.159 | 0.014 | -0.349 | 0.074 | -0.118 | 0.516 |
| 0.47 | 78.835 | 83.780 | -0.119 | 0.061 | -0.314 | 0.105 | -0.140 | 0.438 |
| 0.48 | 75.930 | 84.324 | -0.125 | 0.048 | -0.356 | 0.063 | -0.137 | 0.445 |
| 0.49 | 77.789 | 82.184 | -0.138 | 0.027 | -0.338 | 0.071 | -0.137 | 0.441 |
| 0.50 | 77.361 | 83.422 | -0.072 | 0.261 | -0.270 | 0.166 | -0.122 | 0.497 |

*Note for Table T2:* percent balance improvement 1 = matching of depressed group with current symptoms to depressed group without current symptoms; percent balance improvement 2 = matching of depressed group without current symptoms (matched subgroup) to healthy group; $b_1/p_1$ = unstandardized Betas and *p* values for the comparison of healthy group and depressed group without current symptoms, dependent variable is global FA component (based on all fiber tracts); $b_2/p_2$ = unstandardized Betas and *p* values for the comparison of healthy group and depressed group with current symptoms, dependent variable is global FA component (based on all fiber tracts); $b_3/p_3$ = unstandardized Betas and *p* values for the comparison of the two depressed groups, dependent variable is global FA component (based on all fiber tracts)

### 2. Supplementary Results

#### 2.1 Descriptive statistics for fiber tracts and loadings on principal components

**Table T3** Descriptive statistics for individual fiber tracts (all participants)

| Fiber tract | Healthy participants | | | | Depressed participants without current symptoms | | | | Depressed participants with current symptoms | | | |
|---|---|---|---|---|---|---|---|---|---|---|---|---|
| | *M* | *SD* | Median | Range | *M* | *SD* | Median | Range | *M* | *SD* | Median | Range |
| Acoustic radiation (left) | 0.423 | 0.022 | 0.423 | 0.364-0.483 | 0.423 | 0.021 | 0.423 | 0.365-0.483 | 0.423 | 0.020 | 0.422 | 0.373-0.483 |
| Acoustic radiation (right) | 0.411 | 0.021 | 0.411 | 0.354-0.469 | 0.409 | 0.020 | 0.409 | 0.354-0.469 | 0.410 | 0.020 | 0.411 | 0.356-0.469 |
| Anterior thalamic radiation (left) | 0.398 | 0.018 | 0.398 | 0.349-0.446 | 0.398 | 0.018 | 0.398 | 0.350-0.446 | 0.399 | 0.017 | 0.400 | 0.355-0.446 |
| Anterior thalamic radiation (right) | 0.391 | 0.017 | 0.391 | 0.343-0.438 | 0.390 | 0.017 | 0.391 | 0.344-0.438 | 0.391 | 0.017 | 0.390 | 0.345-0.438 |
| Cingulate gyrus part of cingulum (left) | 0.533 | 0.033 | 0.534 | 0.441-0.625 | 0.531 | 0.033 | 0.532 | 0.440-0.625 | 0.533 | 0.034 | 0.534 | 0.439-0.625 |
| Cingulate gyrus part of cingulum (right) | 0.495 | 0.033 | 0.495 | 0.404-0.586 | 0.494 | 0.032 | 0.494 | 0.407-0.586 | 0.497 | 0.030 | 0.500 | 0.415-0.586 |
| Parahippocampal part of cingulum (left) | 0.313 | 0.026 | 0.314 | 0.240-0.384 | 0.312 | 0.026 | 0.312 | 0.242-0.384 | 0.312 | 0.028 | 0.313 | 0.237-0.384 |
| Parahippocampal part of cingulum (right) | 0.310 | 0.028 | 0.311 | 0.233-0.387 | 0.309 | 0.028 | 0.310 | 0.232-0.387 | 0.309 | 0.029 | 0.309 | 0.233-0.387 |
| Corticospinal tract (left) | 0.545 | 0.021 | 0.546 | 0.486-0.605 | 0.544 | 0.021 | 0.545 | 0.485-0.605 | 0.543 | 0.023 | 0.545 | 0.479-0.605 |
| Corticospinal tract (right) | 0.537 | 0.022 | 0.538 | 0.475-0.598 | 0.535 | 0.022 | 0.536 | 0.473-0.598 | 0.535 | 0.022 | 0.535 | 0.480-0.598 |
| Forceps major | 0.583 | 0.025 | 0.584 | 0.513-0.651 | 0.582 | 0.025 | 0.584 | 0.514-0.651 | 0.581 | 0.027 | 0.583 | 0.513-0.651 |
| Forceps minor | 0.463 | 0.020 | 0.463 | 0.407-0.518 | 0.463 | 0.020 | 0.464 | 0.408-0.518 | 0.464 | 0.020 | 0.464 | 0.409-0.518 |
| Inferior fronto-occipital fasciculus (left) | 0.477 | 0.021 | 0.477 | 0.418-0.535 | 0.478 | 0.020 | 0.478 | 0.422-0.535 | 0.479 | 0.021 | 0.481 | 0.420-0.535 |
| Inferior fronto-occipital fasciculus (right) | 0.464 | 0.020 | 0.464 | 0.409-0.519 | 0.464 | 0.019 | 0.465 | 0.411-0.519 | 0.464 | 0.020 | 0.464 | 0.408-0.519 |
| Inferior longitudinal fasciculus (left) | 0.461 | 0.020 | 0.462 | 0.407-0.514 | 0.462 | 0.019 | 0.463 | 0.409-0.514 | 0.461 | 0.021 | 0.462 | 0.406-0.514 |
| Inferior longitudinal fasciculus (right) | 0.450 | 0.019 | 0.451 | 0.398-0.501 | 0.451 | 0.018 | 0.451 | 0.400-0.501 | 0.450 | 0.019 | 0.451 | 0.398-0.501 |
| Middle cerebellar peduncle | 0.482 | 0.026 | 0.482 | 0.411-0.554 | 0.481 | 0.026 | 0.481 | 0.409-0.554 | 0.481 | 0.024 | 0.480 | 0.409-0.554 |
| Medial lemniscus (left) | 0.425 | 0.024 | 0.425 | 0.360-0.491 | 0.424 | 0.023 | 0.423 | 0.360-0.491 | 0.423 | 0.023 | 0.422 | 0.366-0.491 |

| | | | | | | | | | | | | |
|---|---|---|---|---|---|---|---|---|---|---|---|---|
| Medial lemniscus (right) | 0.426 | 0.024 | 0.426 | 0.360-0.493 | 0.424 | 0.023 | 0.424 | 0.360-0.493 | 0.421 | 0.024 | 0.419 | 0.365-0.493 |
| Posterior thalamic radiation (left) | 0.460 | 0.020 | 0.461 | 0.403-0.516 | 0.460 | 0.020 | 0.461 | 0.406-0.516 | 0.459 | 0.021 | 0.461 | 0.403-0.516 |
| Posterior thalamic radiation (right) | 0.455 | 0.019 | 0.455 | 0.401-0.509 | 0.455 | 0.019 | 0.455 | 0.400-0.509 | 0.453 | 0.021 | 0.454 | 0.393-0.509 |
| Superior longitudinal fasciculus (left) | 0.444 | 0.020 | 0.445 | 0.388-0.499 | 0.445 | 0.020 | 0.446 | 0.388-0.499 | 0.445 | 0.021 | 0.446 | 0.391-0.499 |
| Superior longitudinal fasciculus (right) | 0.425 | 0.020 | 0.425 | 0.370-0.479 | 0.425 | 0.019 | 0.426 | 0.372-0.479 | 0.426 | 0.019 | 0.425 | 0.375-0.479 |
| Superior thalamic radiation (left) | 0.424 | 0.018 | 0.423 | 0.374-0.473 | 0.422 | 0.017 | 0.422 | 0.375-0.473 | 0.421 | 0.018 | 0.423 | 0.375-0.473 |
| Superior thalamic radiation (right) | 0.421 | 0.018 | 0.421 | 0.369-0.472 | 0.419 | 0.018 | 0.419 | 0.371-0.472 | 0.419 | 0.018 | 0.418 | 0.370-0.472 |
| Uncinate fasciculus (left) | 0.386 | 0.023 | 0.386 | 0.323-0.449 | 0.387 | 0.022 | 0.387 | 0.324-0.449 | 0.388 | 0.024 | 0.389 | 0.329-0.449 |
| Uncinate fasciulus (right) | 0.385 | 0.020 | 0.386 | 0.330-0.441 | 0.386 | 0.020 | 0.386 | 0.330-0.441 | 0.386 | 0.021 | 0.385 | 0.334-0.441 |

**Table T4** Descriptive statistics for individual fiber tracts (matched subgroups)

| Fiber tract | Healthy participants | | | | Depressed participants without current symptoms | | | | Depressed participants with current symptoms | | | |
|---|---|---|---|---|---|---|---|---|---|---|---|---|
| | *M* | *SD* | Median | Range | *M* | *SD* | Median | Range | *M* | *SD* | Median | Range |
| Acoustic radiation (left) | 0.423 | 0.021 | 0.423 | 0.367-0.479 | 0.423 | 0.021 | 0.423 | 0.366-0.479 | 0.423 | 0.021 | 0.422 | 0.373-0.479 |
| Acoustic radiation (right) | 0.410 | 0.020 | 0.410 | 0.353-0.467 | 0.409 | 0.020 | 0.409 | 0.355-0.467 | 0.410 | 0.020 | 0.411 | 0.356-0.467 |
| Anterior thalamic radiation (left) | 0.400 | 0.017 | 0.400 | 0.353-0.447 | 0.399 | 0.017 | 0.399 | 0.351-0.447 | 0.399 | 0.017 | 0.400 | 0.355-0.447 |
| Anterior thalamic radiation (right) | 0.392 | 0.017 | 0.392 | 0.346-0.438 | 0.391 | 0.017 | 0.392 | 0.346-0.438 | 0.391 | 0.017 | 0.390 | 0.345-0.438 |
| Cingulate gyrus part of cingulum (left) | 0.534 | 0.033 | 0.536 | 0.443-0.624 | 0.532 | 0.032 | 0.533 | 0.443-0.624 | 0.532 | 0.034 | 0.534 | 0.439-0.624 |
| Cingulate gyrus part of cingulum (right) | 0.495 | 0.033 | 0.495 | 0.404-0.586 | 0.495 | 0.032 | 0.495 | 0.408-0.586 | 0.497 | 0.031 | 0.500 | 0.414-0.586 |
| Parahippocampal part of cingulum (left) | 0.313 | 0.026 | 0.314 | 0.241-0.381 | 0.312 | 0.026 | 0.312 | 0.241-0.381 | 0.312 | 0.028 | 0.313 | 0.237-0.381 |
| Parahippocampal part of cingulum (right) | 0.309 | 0.027 | 0.310 | 0.235-0.381 | 0.309 | 0.028 | 0.309 | 0.232-0.381 | 0.309 | 0.029 | 0.309 | 0.236-0.381 |
| Corticospinal tract (left) | 0.544 | 0.021 | 0.545 | 0.485-0.603 | 0.544 | 0.021 | 0.545 | 0.485-0.603 | 0.543 | 0.023 | 0.544 | 0.479-0.603 |
| Corticospinal tract (right) | 0.535 | 0.022 | 0.536 | 0.473-0.597 | 0.535 | 0.022 | 0.536 | 0.472-0.597 | 0.534 | 0.022 | 0.535 | 0.480-0.597 |
| Forceps major | 0.584 | 0.024 | 0.584 | 0.518-0.646 | 0.583 | 0.024 | 0.584 | 0.516-0.646 | 0.581 | 0.027 | 0.584 | 0.513-0.646 |
| Forceps minor | 0.465 | 0.019 | 0.466 | 0.413-0.517 | 0.464 | 0.020 | 0.465 | 0.410-0.517 | 0.464 | 0.020 | 0.464 | 0.409-0.517 |
| Inferior fronto-occipital fasciculus (left) | 0.480 | 0.020 | 0.480 | 0.422-0.536 | 0.479 | 0.020 | 0.479 | 0.423-0.536 | 0.479 | 0.021 | 0.481 | 0.420-0.536 |
| Inferior fronto-occipital fasciculus (right) | 0.466 | 0.019 | 0.467 | 0.413-0.518 | 0.465 | 0.019 | 0.465 | 0.412-0.518 | 0.464 | 0.021 | 0.464 | 0.408-0.518 |
| Inferior longitudinal fasciculus (left) | 0.464 | 0.019 | 0.465 | 0.413-0.515 | 0.463 | 0.019 | 0.464 | 0.412-0.515 | 0.461 | 0.021 | 0.462 | 0.406-0.515 |
| Inferior longitudinal fasciculus (right) | 0.453 | 0.018 | 0.453 | 0.403-0.501 | 0.451 | 0.018 | 0.452 | 0.401-0.501 | 0.450 | 0.019 | 0.451 | 0.398-0.501 |
| Middle cerebellar peduncle | 0.481 | 0.025 | 0.481 | 0.412-0.548 | 0.481 | 0.025 | 0.481 | 0.409-0.548 | 0.481 | 0.024 | 0.480 | 0.409-0.548 |
| Medial lemniscus (left) | 0.423 | 0.023 | 0.423 | 0.359-0.487 | 0.423 | 0.023 | 0.423 | 0.361-0.487 | 0.423 | 0.023 | 0.422 | 0.366-0.487 |
| Medial lemniscus (right) | 0.424 | 0.023 | 0.424 | 0.362-0.487 | 0.424 | 0.023 | 0.423 | 0.360-0.487 | 0.421 | 0.024 | 0.419 | 0.365-0.487 |
| Posterior thalamic radiation (left) | 0.463 | 0.019 | 0.464 | 0.412-0.515 | 0.461 | 0.019 | 0.462 | 0.408-0.515 | 0.459 | 0.021 | 0.461 | 0.403-0.515 |
| Posterior thalamic radiation (right) | 0.457 | 0.018 | 0.457 | 0.406-0.508 | 0.455 | 0.019 | 0.456 | 0.403-0.508 | 0.453 | 0.021 | 0.454 | 0.393-0.508 |
| Superior longitudinal fasciculus (left) | 0.446 | 0.019 | 0.447 | 0.394-0.499 | 0.446 | 0.020 | 0.446 | 0.389-0.499 | 0.445 | 0.021 | 0.446 | 0.391-0.499 |

| | | | | | | | | | | | | |
|---|---|---|---|---|---|---|---|---|---|---|---|---|
| Superior longitudinal fasciculus (right) | 0.426 | 0.019 | 0.427 | 0.374-0.479 | 0.426 | 0.019 | 0.427 | 0.373-0.479 | 0.426 | 0.019 | 0.426 | 0.375-0.479 |
| Superior thalamic radiation (left) | 0.423 | 0.017 | 0.422 | 0.377-0.469 | 0.422 | 0.017 | 0.421 | 0.376-0.469 | 0.421 | 0.018 | 0.422 | 0.375-0.469 |
| Superior thalamic radiation (right) | 0.420 | 0.017 | 0.420 | 0.374-0.466 | 0.419 | 0.017 | 0.419 | 0.371-0.466 | 0.419 | 0.018 | 0.418 | 0.370-0.466 |
| Uncinate fasciculus (left) | 0.388 | 0.022 | 0.388 | 0.327-0.451 | 0.387 | 0.022 | 0.387 | 0.327-0.451 | 0.388 | 0.024 | 0.388 | 0.329-0.451 |
| Uncinate fasciculus (right) | 0.387 | 0.020 | 0.388 | 0.355-0.440 | 0.386 | 0.020 | 0.386 | 0.330-0.44 | 0.386 | 0.021 | 0.385 | 0.334-0.44 |

**Table T5** Loadings of individual fiber tracts on the four PCs (for all participants)

| Tract | All | Association/ commissural fibers | Thalamic radiations | Projection fibers |
|---|---|---|---|---|
| Cingulate gyrus part of cingulum (left) | -0.1504 | -0.2044 | | |
| Cingulate gyrus part of cingulum (right) | -0.1443 | -0.1972 | | |
| Inferior fronto-occipital fasciculus (left) | -0.2452 | -0.3256 | | |
| Inferior fronto-occipital fasciculus (right) | -0.2519 | -0.33 | | |
| Inferior longitudinal fasciculus (left) | -0.2385 | -0.3211 | | |
| Inferior longitudinal fasciculus (right) | -0.2453 | -0.3257 | | |
| Parahippocampal part of cingulum (left) | -0.1048 | -0.1385 | | |
| Parahippocampal part of cingulum (right) | -0.0916 | -0.1185 | | |
| Superior longitudinal fasciculus (left) | -0.2203 | -0.2976 | | |
| Superior longitudinal fasciculus (right) | -0.2297 | -0.3039 | | |
| Uncinate fasciculus (left) | -0.1918 | -0.26 | | |
| Uncinate fasciculus (right) | -0.1964 | -0.2674 | | |
| Forceps major | -0.1699 | -0.221 | | |
| Forceps minor | -0.2319 | -0.3052 | | |
| Anterior thalamic radiation (left) | -0.2323 | | -0.4331 | |
| Anterior thalamic radiation (right) | -0.2331 | | -0.4394 | |
| Posterior thalamic radiation (left) | -0.2069 | | -0.3802 | |
| Posterior thalamic radiation (right) | -0.2182 | | -0.4038 | |
| Superior thalamic radiation (left) | -0.195 | | -0.3908 | |
| Superior thalamic radiation (right) | -0.1972 | | -0.3988 | |
| Acoustic radiation (left) | -0.1876 | | | -0.393 |
| Acoustic radiation (right) | -0.1853 | | | -0.4022 |
| Corticospinal tract (left) | -0.1859 | | | -0.4765 |
| Corticospinal tract (right) | -0.1842 | | | -0.4753 |
| Medial lemniscus (left) | -0.0727 | | | -0.2723 |
| Medial lemniscus (right) | -0.0753 | | | -0.275 |
| Middle cerebellar peduncle | -0.1126 | | | -0.2846 |

*Note.* Values represent loadings of individual fiber tracts on the first principal components from the PCAs. Scores on principal components were sign-adjusted for the analyses, so higher values represent higher fiber tract integrity.

**Table T6** Loadings of individual fiber tracts on the four PCs (for matched subsample)

| Tract | All | Association/ commissural fibers | Thalamic radiation | Projection fibers |
|---|---|---|---|---|
| Cingulate gyrus part of cingulum (left) | 0.1431 | -0.1977 | | |
| Cingulate gyrus part of cingulum (right) | 0.1367 | -0.1915 | | |
| Inferior fronto-occipital fasciculus (left) | 0.2487 | -0.3296 | | |
| Inferior fronto-occipital fasciculus (right) | 0.2546 | -0.3325 | | |
| Inferior longitudinal fasciculus (left) | 0.2393 | -0.3219 | | |
| Inferior longitudinal fasciculus (right) | 0.2443 | -0.3252 | | |
| Parahippocampal part of cingulum (left) | 0.1020 | -0.1341 | | |
| Parahippocampal part of cingulum (right) | 0.0901 | -0.1175 | | |
| Superior longitudinal fasciculus (left) | 0.2202 | -0.3001 | | |
| Superior longitudinal fasciculus (right) | 0.2309 | -0.3075 | | |
| Uncinate fasciculus (left) | 0.1896 | -0.2579 | | |
| Uncinate fasciculus (right) | 0.197 | -0.2684 | | |
| Forceps major | 0.1658 | -0.2186 | | |
| Forceps minor | 0.2311 | -0.305 | | |
| Anterior thalamic radiation (left) | 0.2336 | | -0.4299 | |
| Anterior thalamic radiation (right) | 0.2351 | | -0.4371 | |
| Posterior thalamic radiation (left) | 0.2087 | | -0.3893 | |
| Posterior thalamic radiation (right) | 0.2168 | | -0.4054 | |
| Superior thalamic radiation (left) | 0.1971 | | -0.3859 | |
| Superior thalamic radiation (right) | 0.2007 | | -0.3991 | |
| Acoustic radiation (left) | 0.1858 | | | 0.3821 |
| Acoustic radiation (right) | 0.1858 | | | 0.3986 |
| Corticospinal tract (left) | 0.1841 | | | 0.4749 |
| Corticospinal tract (right) | 0.1805 | | | 0.4747 |
| Medial lemniscus (left) | 0.0776 | | | 0.2772 |
| Medial lemniscus (right) | 0.0794 | | | 0.2811 |
| Middle cerebellar peduncle | 0.1179 | | | 0.2973 |

*Note.* Values represent loadings of individual fiber tracts on the first principal components from the PCAs. Scores on principal components for association/commissural fibers and thalamic fibers were sign-adjusted for the analyses, so higher values represent higher fiber tract integrity.

### 2.2 Multiple regression analyses for global FA measures

**Figure S1** Scree plots for the principal component analyses

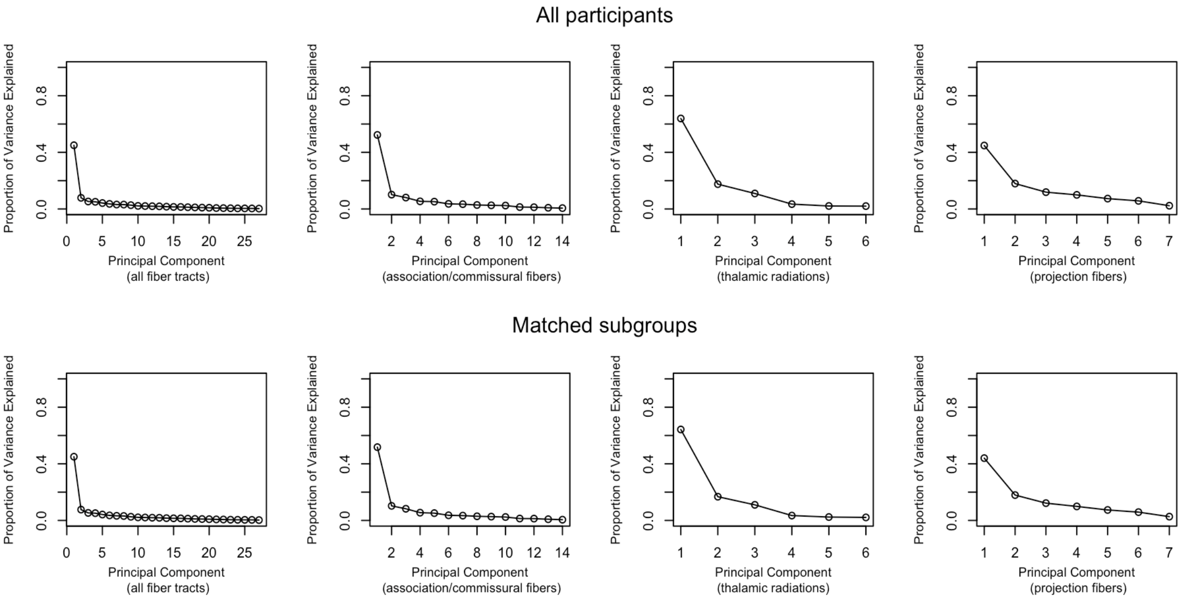


*Note.* The upper four plots depict the explained variance of the PCAs including all participants, the lower four the explained variance of the PCAs including the matched subsample (propensity score matching, method: *nearest neighbor*, caliper = 0.05).

**Table T7** Multiple regression analyses with global FA components as dependent variables (healthy vs. depressed without current symptoms; all participants)

DV: integrity of all fiber tracts

| | ***b*** | ***SE*** | ***t* value** |
|---|---|---|---|
| Depression status | -0.137 | 0.05 | -2.738* |
| Age | 0.069 | 0.046 | 1.479 |
| Age$^2$ | -0.001 | 0 | -3.469* |
| Sex: Male | 0.518 | 0.031 | 16.496* |
| Image Quality Rating | -2.18 | 0.148 | -14.715* |
| Education status | 0.047 | 0.018 | 2.691* |
| Antidepressant intake: true | -0.152 | 0.112 | -1.354 |
| Assessment center 1 | 0.716 | 0.066 | 10.841* |
| Assessment center 2 | -0.267 | 0.051 | -5.207* |
| $R^2$ overall | 0.088* | | |
| $F$(9,18663)* | 200.9 | | |

DV: integrity of association/commissural fibers

| | ***b*** | ***SE*** | ***t* value** |
|---|---|---|---|
| Depression status | -0.096 | 0.04 | -2.398* |
| Age | 0.118 | 0.037 | 3.189* |
| Age$^2$ | -0.002 | 0 | -5.628* |
| Sex: Male | 0.229 | 0.025 | 9.162* |
| Image Quality Rating | -1.333 | 0.118 | -11.313* |
| Education status | 0.022 | 0.014 | 1.560 |
| Antidepressant intake: true | -0.117 | 0.089 | -1.314 |
| Assessment center 1 | 0.369 | 0.053 | 7.024* |
| Assessment center 2 | -0.268 | 0.041 | -6.565* |
| $R^2$ overall | 0.097* | | |
| $F$(9,18694)* | 224.3 | | |

DV: integrity of thalamic radiations

| | ***b*** | ***SE*** | ***t* value** |
|---|---|---|---|
| Depression status | -0.101 | 0.028 | -3.807* |
| Age | 0.024 | 0.026 | 0.918 |
| Age$^2$ | -0.001 | 0 | -2.649* |
| Sex: Male | 0.245 | 0.017 | 14.088* |
| Image Quality Rating | -1.345 | 0.082 | -16.360* |
| Education status | 0.034 | 0.01 | 3.463* |
| Antidepressant intake: true | -0.149 | 0.089 | -1.314 |
| Assessment center 1 | 0.598 | 0.037 | 16.348* |
| Assessment center 2 | -0.006 | 0.028 | -0.212 |
| $R^2$ overall | 0.081* | | |
| $F$(9,18622)* | 183.5 | | |

DV: integrity of projection fibers

| | *b* | *SE* | *t* value |
|---|---|---|---|
| Depression status | -0.02 | 0.024 | -0.841 |
| Age | -0.06 | 0.022 | -2.705* |
| Age$^2$ | 0.000 | 0 | 2.533* |
| Sex: Male | 0.58 | 0.015 | 38.454* |
| Image Quality Rating | -1.319 | 0.071 | -18.524* |
| Education status | 0.035 | 0.008 | 4.208* |
| Antidepressant intake: true | -0.007 | 0.054 | -0.125 |
| Assessment center 1 | 0.357 | 0.032 | 11.258* |
| Assessment center 2 | -0.11 | 0.025 | -4.464 |
| $R^2$ overall | 0.098* | | |
| $F$(9,18648)* | 226.4 | | |

*Note*. DV = dependent variable; Depression status: 0 = healthy, 1 = depressed without current symptoms; sex: 0 = female, 1 = male; Education status: 0 = incomplete, 1 = compulsory, 2 = continued, 3 = college, 4 = university; Assessment center is dummy-coded. *$p$ < .05.

**Table T8** Multiple regression analyses with global FA components as dependent variables (healthy vs. depressed with current symptoms; all participants)

DV: integrity of all fiber tracts

| | *b* | *SE* | *t* value |
|---|---|---|---|
| Depression status | -0.303 | 0.188 | -1.617 |
| Age | 0.059 | 0.055 | 1.083 |
| Age$^2$ | -0.001 | 0 | -2.761* |
| Sex: Male | 0.518 | 0.037 | 14.038* |
| Image Quality Rating | -2.28 | 0.171 | -13.331* |
| Education status | 0.052 | 0.02 | 2.564* |
| Antidepressant intake: true | 0.196 | 0.226 | 0.863 |
| Assessment center 1 | 0.781 | 0.077 | 10.197* |
| Assessment center 2 | -0.219 | 0.061 | -3.609* |
| $R^2$ overall | 0.092 | | |
| $F$(9, 13473)* | 151.1 | | |

DV integrity of association/commissural fibers

| | *b* | *SE* | *t* value |
|---|---|---|---|
| Depression status | -0.308 | 0.15 | -2.060* |
| Age | 0.108 | 0.043 | 2.495* |
| Age$^2$ | -0.002 | 0 | -4.593* |
| Sex: Male | 0.229 | 0.029 | 7.814* |
| Image Quality Rating | -1.399 | 0.136 | -10.296* |
| Education status | 0.029 | 0.016 | 1.791 |
| Antidepressant intake: true | 0.201 | 0.179 | 1.118 |
| Assessment center 1 | 0.394 | 0.061 | 6.456* |
| Assessment center 2 | -0.241 | 0.0484 | -4.991* |
| $R^2$ overall | 0.102 | | |
| $F$(9, 13486)* | 170.6 | | |

DV integrity of thalamic radiations

| | b | *SE* | *t* value |
|---|---|---|---|
| Depression status | -0.181 | 0.104 | -1.735 |
| Age | 0.025 | 0.03 | 0.830 |
| Age$^2$ | -0.001 | 0 | -2.238* |
| Sex: Male | 0.242 | 0.02 | 11.865* |
| Image Quality Rating | -1.408 | 0.095 | -14.842* |
| Education status | 0.037 | 0.011 | 3.244* |
| Antidepressant intake: true | -0.016 | 0.125 | -0.128 |
| Assessment center 1 | 0.624 | 0.042 | 14.692* |
| Assessment center 2 | 0.011 | 0.034 | 0.335 |
| $R^2$ overall | 0.082 | | |
| $F$(9, 13440)* | 135.1 | | |

DV integrity of projection fibers

| | b | *SE* | *t* value |
|---|---|---|---|
| Depression status | 0.003 | 0.09 | 0.035 |
| Age | -0.059 | 0.026 | -2.230* |
| Age$^2$ | < 0.001 | 0 | 2.168* |
| Sex: Male | 0.578 | 0.018 | 32.444* |
| Image Quality Rating | -1.379 | 0.082 | -16.724* |
| Education status | 0.031 | 0.01 | 3.210* |
| Antidepressant intake: true | -0.099 | 0.109 | -0.910 |
| Assessment center 1 | 0.391 | 0.037 | 10.546* |
| Assessment center 2 | -0.096 | 0.029 | -3.288* |
| $R^2$ overall | 0.099 | | |
| $F$(9, 13478)* | 164.5 | | |

*Note*. DV = dependent variable; Depression status: 0 = healthy, 1 = depressed without current symptoms; sex: 0 = female, 1 = male; Education status: 0 = incomplete, 1 = compulsory, 2 = continued, 3 = college, 4 = university; Assessment center is dummy-coded. *$p < .05$.

**Table T9** Multiple regression analyses with global FA components as dependent variables (depressed with current symptoms vs. depressed without current symptoms; all participants)

DV: integrity of all fiber tracts

| | *b* | *SE* | *t* value |
|---|---|---|---|
| Depression status | -0.101 | 0.18 | -0.564 |
| Age | 0.119 | 0.086 | 1.376 |
| Age$^2$ | -0.002 | 0.001 | -2.540* |
| Sex: Male | 0.555 | 0.057 | 9.734* |
| Image Quality Rating | -1.68 | 0.278 | -6.032* |
| Education status | 0.016 | 0.033 | 0.492 |
| Antidepressant intake: true | -0.243 | 0.117 | -2.08* |
| Assessment center 1 | 0.717 | 0.123 | 5.822* |
| Assessment center 2 | -0.36 | 0.091 | -3.942* |
| $R^2$ overall | 0.088 | | |
| $F$(9, 5750)* | 61.67 | | |

DV: integrity of association/commissural fibers

| | *b* | *SE* | *t* value |
|---|---|---|---|
| Depression status | -0.143 | 0.144 | -0.995 |
| Age | 0.156 | 0.069 | 2.255* |
| Age$^2$ | -0.002 | 0.001 | -3.579* |
| Sex: Male | 0.256 | 0.046 | 5.628* |
| Image Quality Rating | -0.998 | 0.222 | -4.501* |
| Education status | -0.009 | 0.026 | -0.329 |
| Antidepressant intake: true | -0.155 | 0.093 | -1.664 |
| Assessment center 1 | 0.379 | 0.098 | 3.867* |
| Assessment center 2 | -0.316 | 0.073 | -4.338* |
| $R^2$ overall | 0.09 | | |
| $F(9, 5766)$* | 63.62 | | |

DV: integrity of thalamic radiations

| | *b* | *SE* | *t* value |
|---|---|---|---|
| Depression status | -0.069 | 0.01 | -0.695 |
| Age | 0.059 | 0.048 | 1.233 |
| Age$^2$ | -0.001 | 0 | -2.335 |
| Sex: Male | 0.271 | 0.032 | 8.567* |
| Image Quality Rating | -1.086 | 0.154 | -7.048* |
| Education status | 0.02 | 0.018 | 1.106 |
| Antidepressant intake: true | -0.172 | 0.065 | -2.659* |
| Assessment center 1 | 0.62 | 0.068 | 9.084* |
| Assessment center 2 | -0.045 | 0.051 | -0.895 |
| $R^2$ overall | 0.086 | | |
| $F(9, 5736)$* | 60.27 | | |

DV: integrity of projection fibers

| | *b* | *SE* | *t* value |
|---|---|---|---|
| Depression status | -0.021 | 0.085 | -0.247 |
| Age | -0.056 | 0.041 | -1.351 |
| Age$^2$ | < 0.001 | 0 | 1.025 |
| Sex: Male | 0.575 | 0.027 | 21.153* |
| Image Quality Rating | -0.985 | 0.133 | -7.405* |
| Education status | 0.042 | 0.016 | 2.709* |
| Antidepressant intake: true | -0.035 | 0.055 | -0.605 |
| Assessment center 1 | 0.352 | 0.058 | 6.032* |
| Assessment center 2 | -0.133 | 0.044 | -3.058* |
| $R^2$ overall | 0.093 | | |
| $F(9, 5732)$* | 65.07 | | |

*Note*. DV = dependent variable; Depression status: 0 = healthy, 1 = depressed without current symptoms; sex: 0 = female, 1 = male; Education status: 0 = incomplete, 1 = compulsory, 2 = continued, 3 = college, 4 = university; Assessment center is dummy-coded. $*p < .05$.

**Table T10** Multiple regression analyses with global FA components as dependent variables (healthy vs. depressed without current symptoms; matched subgroups)

DV: integrity of all fiber tracts

| | ***b*** | ***SE*** | ***t* value** |
|---|---|---|---|
| Depression status | -0.184 | 0.069 | -2.673* |
| Age | 0.114 | 0.079 | 1.450 |
| $Age^2$ | -0.002 | 0.001 | -2.592* |
| Sex: Male | 0.453 | 0.05 | 8.999* |
| Image Quality Rating | -2.413 | 0.252 | -9.589* |
| Education status | 0.009 | 0.028 | 0.315 |
| Antidepressant intake: true | 0.141 | 0.14 | -1.012 |
| Assessment center 1 | 0.797 | 0.107 | 7.481* |
| Assessment center 2 | -0.193 | 0.079 | -2.438* |
| $R^2$ overall | 0.077* | | |
| $F(9, 8277)$* | 76.76 | | |

DV integrity of association/commissural fibers

| | ***b*** | ***SE*** | ***t* value** |
|---|---|---|---|
| Depression status | -0.129 | 0.055 | -2.372* |
| Age | 0.11 | 0.062 | 1.764 |
| $Age^2$ | -0.002 | 0.001 | -3.079* |
| Sex: Male | 0.17 | 0.04 | 4.273* |
| Image Quality Rating | -1.477 | 0.199 | -7.427* |
| Education status | 0.002 | -0.002 | 0.087 |
| Antidepressant intake: true | -0.17 | 0.11 | -1.545 |
| Assessment center 1 | 0.431 | 0.084 | 5.124* |
| Assessment center 2 | -0.189 | 0.063 | -3.013* |
| $R^2$ overall | 0.077* | | |
| $F(9, 8278)$* | 76.35 | | |

DV integrity of thalamic fibers

| | ***b*** | ***SE*** | ***t* value** |
|---|---|---|---|
| Depression status | -0.132 | 0.038 | -3.480 |
| Age | 0.052 | 0.043 | 1.209 |
| $Age^2$ | -0.001 | 0 | -2.264 |
| Sex: Male | 0.202 | 0.028 | 7.288 |
| Image Quality Rating | -1.384 | 0.139 | -9.980 |
| Education status | 0.012 | 0.016 | 0.774 |
| Antidepressant intake: true | -0.136 | 0.077 | -1.781 |
| Assessment center 1 | 0.65 | 0.059 | 11.081 |
| Assessment center 2 | 0.026 | 0.0435 | 0.604 |
| $R^2$ overall | 0.075* | | |
| $F(9, 8247)$* | 74.67 | | |

DV: integrity of projection fibers

| | *b* | *SE* | *t* value |
|---|---|---|---|
| Depression status | -0.015 | 0.033 | -0.448 |
| Age | -0.052 | 0.038 | -1.350 |
| Age$^2$ | <0.001 | 0 | 1.158 |
| Sex: Male | 0.563 | 0.024 | 23.123 |
| Image Quality Rating | -1.466 | 0.122 | -12.033 |
| Education status | 0.019 | 0.014 | 1.359 |
| Antidepressant intake: true | 0.02 | 0.067 | 0.299 |
| Assessment center 1 | 0.383 | 0.051 | 7.416 |
| Assessment center 2 | -0.111 | 0.038 | -2.899 |
| $R^2$ overall | 0.083* | | |
| $F$(9, 8263)* | 82.85 | | |

*Note*. DV = dependent variable; Depression status: 0 = healthy, 1 = depressed without current symptoms; sex: 0 = female, 1 = male; Education status: 0 = incomplete, 1 = compulsory, 2 = continued, 3 = college, 4 = university; Assessment center is dummy-coded. *$p$ < .05.

**Table T11** Multiple regression analyses with PCs as dependent variables (healthy vs. depressed with current symptoms; matched subgroups)

DV: integrity of all fiber tracts

| | *b* | *SE* | *t* value |
|---|---|---|---|
| Depression status | -0.372 | 0.203 | -1.830 |
| Age | 0.053 | 0.107 | 0.491 |
| Age$^2$ | -0.001 | 0.001 | -1.266 |
| Sex: Male | 0.439 | 0.069 | 6.368‘ |
| Image Quality Rating | -2.716 | 0.34 | -7.990* |
| Education status | 0.01 | 0.038 | 0.274 |
| Antidepressant intake: true | 0.191 | 0.314 | 0.610 |
| Assessment center 1 | 1.065 | 0.143 | 7.435* |
| Assessment center 2 | -0.035 | 0.109 | -0.322 |
| $R^2$ overall | 0.078* | | |
| $F$(9, 4418)* | 41.63 | | |

DV: integrity of association/commissural fibers

| | *b* | *SE* | *t* value |
|---|---|---|---|
| Depression status | -0.38 | 0.161 | -2.356* |
| Age | 0.066 | 0.085 | 0.781 |
| Age$^2$ | -0.001 | 0.001 | -1.683 |
| Sex: Male | 0.156 | 0.055 | 2.866* |
| Image Quality Rating | -1.715 | 0.269 | -6.383* |
| Education status | 0.023 | 0.03 | 0.765 |
| Antidepressant intake: true | 0.248 | 0.248 | 0.999 |
| Assessment center 1 | 0.546 | 0.113 | 4.816* |
| Assessment center 2 | -0.065 | 0.086 | -0.760 |
| $R^2$ overall | 0.074* | | |
| $F$(9, 4413)* | 39.6 | | |

DV: integrity of thalamic fibers

| | *b* | *SE* | *t* value |
|---|---|---|---|
| Depression status | -0.252 | 0.111 | -2.271* |
| Age | 0.034 | 0.058 | 0.581 |
| Age$^2$ | -0.001 | 0 | -1.282 |
| Sex: Male | 0.191 | 0.038 | 5.089* |
| Image Quality Rating | -1.446 | 0.185 | -7.798* |
| Education status | 0.011 | 0.021 | 0.553 |
| Antidepressant intake: true | 0.058 | 0.171 | 0.340 |
| Assessment center 1 | 0.788 | 0.078 | 10.129* |
| Assessment center 2 | 0.086 | 0.059 | 1.463 |
| $R^2$ overall | 0.076* | | |
| $F$(9, 4397)* | 40.54 | | |

DV: integrity of projection fibers

| | *b* | *SE* | *t* value |
|---|---|---|---|
| Depression status | 0.01 | 0.099 | 0.106 |
| Age | -0.066 | 0.052 | -1.259 |
| Age$^2$ | < 0.001 | 0 | 1.172 |
| Sex: Male | 0.533 | 0.033 | 15.800* |
| Image Quality Rating | -1.634 | 0.167 | -9.804* |
| Education status | 0.009 | 0.018 | 0.500 |
| Antidepressant intake: true | -0.172 | 0.153 | -1.121 |
| Assessment center 1 | 0.505 | 0.07 | 7.181* |
| Assessment center 2 | -0.077 | 0.053 | -1.447 |
| $R^2$ overall | 0.083* | | |
| $F$(9, 4422)* | 44.5 | | |

*Note*. DV = dependent variable; Depression status: 0 = healthy, 1 = depressed without current symptoms; sex: 0 = female, 1 = male; Education status: 0 = incomplete, 1 = compulsory, 2 = continued, 3 = college, 4 = university; Assessment center is dummy-coded. *$p < .05$.

**Table T12** Multiple regression analyses with global FA components as dependent variables (depressed without current symptoms vs. depressed with current symptoms; matched subgroups)

DV: integrity of all fiber tracts

| | *b* | *SE* | *t* value |
|---|---|---|---|
| Depression status | -0.114 | 0.189 | -0.606 |
| Age | 0.195 | 0.109 | 1.796 |
| Age$^2$ | -0.002 | 0.001 | -2.801* |
| Sex: Male | 0.524 | 0.069 | 7.596* |
| Image Quality Rating | -1.689 | 0.343 | -4.922* |
| Education status | -0.006 | 0.039 | -0.159 |
| Antidepressant intake: true | -0.198 | 0.138 | -1.434 |
| Assessment center 1 | 0.757 | 0.147 | 5.115* |
| Assessment center 2 | -0.333 | 0.108 | -3.073* |
| $R^2$ overall | 0.088* | | |
| $F$(9, 4417)* | 47.42 | | |

DV: integrity of association/commissural fibers

| | ***b*** | ***SE*** | ***t* value** |
|---|---|---|---|
| Depression status | -0.165 | 0.149 | -1.101 |
| Age | 0.187 | 0.086 | 2.175* |
| $Age^2$ | -0.002 | 0.001 | -3.293* |
| Sex: Male | 0.227 | 0.055 | 4.157* |
| Image Quality Rating | -0.954 | 0.271 | -3.517* |
| Education status | -0.028 | 0.031 | -0.914 |
| Antidepressant intake: true | -0.172 | 0.109 | -1.585 |
| Assessment center 1 | 0.41 | 0.117 | 3.521* |
| Assessment center 2 | -0.3 | 0.086 | -3.509* |
| $R^2$ overall | 0.089* | | |
| $F(9, 4421)$* | 47.72 | | |

DV: integrity of thalamic radiations

| | ***b*** | ***SE*** | ***t* value** |
|---|---|---|---|
| Depression status | -0.076 | 0.105 | -0.724 |
| Age | 0.095 | 0.061 | 1.570 |
| $Age^2$ | -0.001 | 0 | -2.507 |
| Sex: Male | 0.243 | 0.038 | 6.331 |
| Image Quality Rating | -1.14 | 0.191 | -5.973 |
| Education status | 0.009 | 0.022 | 0.425 |
| Antidepressant intake: true | -0.156 | 0.077 | -2.044 |
| Assessment center 1 | 0.628 | 0.082 | 7.632 |
| Assessment center 2 | -0.036 | 0.06 | -0.596 |
| $R^2$ overall | 0.085* | | |
| $F(9, 4402)$* | 45.57 | | |

DV: integrity of projection fibers

| | ***b*** | ***SE*** | ***t* value** |
|---|---|---|---|
| Depression status | -0.023 | 0.09 | -0.256 |
| Age | -0.054 | 0.052 | -1.029 |
| $Age^2$ | < 0.001 | 0 | 0.713 |
| Sex: Male | 0.581 | 0.033 | 17.531* |
| Image Quality Rating | -1.03 | 0.164 | -6.272* |
| Education status | 0.028 | 0.019 | 1.505 |
| Antidepressant intake: true | -0.01 | 0.066 | -0.155 |
| Assessment center 1 | 0.369 | 0.071 | 5.217* |
| Assessment center 2 | -0.135 | 0.052 | -2.596* |
| $R^2$ overall | 0.086* | | |
| $F(9, 4401)$* | 45.87 | | |

*Note*. DV = dependent variable; Depression status: 0 = healthy, 1 = depressed without current symptoms; sex: 0 = female, 1 = male; Education status: 0 = incomplete, 1 = compulsory, 2 = continued, 3 = college, 4 = university; Assessment center is dummy-coded. $*p < .05$.

### 2.3 Multiple regression analyses for individual fiber tracts

**Table T13** Effects of depression status on FA of individual fiber tracts (analyses with all participants)

| **Fiber tract** | $b_1$ | $p_1$ | $b_2$ | $p_2$ | $b_3$ | $p_3$ |
|---|---|---|---|---|---|---|
| Acoustic radiation (left) | 0.0000 | 0.9128 | 0.0004 | 0.7415 | 0.0002 | 0.8693 |
| Acoustic radiation (right) | -0.0005 | 0.1434 | 0.0003 | 0.8091 | 0.0007 | 0.5382 |
| Anterior thalamic radiation (left) | -0.0006 | 0.0476 | -0.0012 | 0.2574 | 0.0000 | 0.9616 |
| Anterior thalamic radiation (right) | -0.0008 | 0.0062* | -0.0010 | 0.3665 | -0.0001 | 0.8998 |
| Cingulate gyrus part of cingulum (left) | -0.0018 | 0.0008* | -0.0028 | 0.1666 | 0.0000 | 0.9927 |
| Cingulate gyrus part of cingulum (right) | -0.0008 | 0.1543 | 0.0005 | 0.7886 | 0.0024 | 0.2131 |
| Parahippocampal part of cingulum (left) | -0.0003 | 0.4855 | -0.0008 | 0.6340 | -0.0002 | 0.9227 |
| Parahippocampal part of cingulum (right) | -0.0002 | 0.6470 | -0.0002 | 0.9085 | 0.0002 | 0.9191 |
| Corticospinal tract (left) | -0.0002 | 0.4795 | -0.0001 | 0.9150 | -0.0006 | 0.6230 |
| Corticospinal tract (right) | -0.0005 | 0.1771 | 0.0004 | 0.7879 | -0.0002 | 0.8761 |
| Forceps major | -0.0006 | 0.1449 | -0.0020 | 0.2007 | -0.0013 | 0.3833 |
| Forceps minor | -0.0008 | 0.0100* | -0.0017 | 0.1589 | -0.0002 | 0.8283 |
| Inferior fronto-occipital fasciculus (left) | -0.0005 | 0.1413 | -0.0011 | 0.3683 | -0.0002 | 0.8884 |
| Inferior fronto-occipital fasciculus (right) | -0.0008 | 0.0104* | -0.0028 | 0.0197 | -0.0014 | 0.2247 |
| Inferior longitudinal fasciculus (left) | -0.0006 | 0.0634 | -0.0032 | 0.0068* | -0.0025 | 0.0266 |
| Inferior longitudinal fasciculus (right) | -0.0006 | 0.0358 | -0.0034 | 0.0023* | -0.0019 | 0.0782 |
| Middle cerebellar peduncle | 0.0001 | 0.8785 | 0.0003 | 0.8316 | -0.0006 | 0.7096 |
| Medial lemniscus (left) | -0.0002 | 0.6707 | -0.0006 | 0.6617 | -0.0006 | 0.6735 |
| Medial lemniscus (right) | -0.0009 | 0.0173 | -0.0023 | 0.1163 | -0.0025 | 0.0709 |
| Posterior thalamic radiation (left) | -0.0012 | 0.0002* | -0.0036 | 0.0039* | -0.0026 | 0.0261 |
| Posterior thalamic radiation (right) | -0.0014 | 0.0000* | -0.0039 | 0.0009* | -0.0022 | 0.0530 |
| Superior longitudinal fasciculus (left) | -0.0002 | 0.5333 | -0.0016 | 0.1813 | -0.0013 | 0.2854 |
| Superior longitudinal fasciculus (right) | -0.0006 | 0.0787 | -0.0009 | 0.4280 | 0.0000 | 0.9665 |
| Superior thalamic radiation (left) | -0.0007 | 0.0104* | -0.0001 | 0.8987 | -0.0001 | 0.9451 |
| Superior thalamic radiation (right) | -0.0008 | 0.0076* | 0.0001 | 0.9640 | 0.0005 | 0.6182 |
| Uncinate fasciculus (left) | -0.0004 | 0.2842 | 0.0005 | 0.7179 | 0.0015 | 0.2596 |
| Uncinate fasciculus (right) | -0.0006 | 0.0575 | -0.0015 | 0.2141 | 0.0000 | 0.9897 |

*Note.* $b_1/p_1$: Unstandardized Betas and *p* values for depression status derived from multiple regression analyses in the comparison of healthy and depressed participants without current symptoms; $b_1/p_1$: Unstandardized Betas and *p* values for depression status derived from multiple regression analyses in the comparison of healthy and depressed participants with current symptoms; $b_1/p_1$: Unstandardized Betas and *p* values for depression status derived from multiple regression analyses in the comparison of depressed participants with and without current symptoms. *Significant after correction for multiple comparisons (Benjamini-Hochberg procedure, false discovery rate: 5 %)

**Table T14** Effects of depression status on FA of individual fiber tracts (analyses with matched subgroups)

| Fiber tract | $b_1$ | $p_1$ | $b_2$ | $p_2$ | $b_3$ | $p_3$ |
|---|---|---|---|---|---|---|
| Acoustic radiation (left) | 0.0003 | 0.5101 | 0.0007 | 0.6165 | 0.0002 | 0.8840 |
| Acoustic radiation (right) | -0.0005 | 0.2450 | 0.0004 | 0.7413 | 0.0007 | 0.5386 |
| Anterior thalamic radiation (left) | -0.0007 | 0.0556 | -0.0016 | 0.1480 | -0.0001 | 0.8880 |
| Anterior thalamic radiation (right) | -0.0007 | 0.0451 | -0.0015 | 0.1588 | -0.0004 | 0.6597 |
| Cingulate gyrus part of cingulum (left) | -0.0021 | 0.0034* | -0.0035 | 0.1006 | -0.0004 | 0.8277 |
| Cingulate gyrus part of cingulum (right) | -0.0002 | 0.7896 | 0.0002 | 0.9315 | 0.0020 | 0.2961 |
| Parahippocampal part of cingulum (left) | -0.0008 | 0.1914 | -0.0017 | 0.3259 | 0.0000 | 0.9804 |
| Parahippocampal part of cingulum (right) | -0.0008 | 0.2107 | -0.0008 | 0.6555 | 0.0005 | 0.7867 |
| Corticospinal tract (left) | -0.0002 | 0.5965 | -0.0002 | 0.8895 | -0.0007 | 0.5740 |
| Corticospinal tract (right) | -0.0004 | 0.4408 | 0.0004 | 0.7961 | -0.0002 | 0.8639 |
| Forceps major | -0.0005 | 0.3143 | -0.0021 | 0.1922 | -0.0013 | 0.3761 |
| Forceps minor | -0.0011 | 0.0073* | -0.0021 | 0.0849 | -0.0003 | 0.7960 |
| Inferior fronto-occipital fasciculus (left) | -0.0006 | 0.2036 | -0.0015 | 0.2519 | -0.0002 | 0.8758 |
| Inferior fronto-occipital fasciculus (right) | -0.0012 | 0.0034* | -0.0034 | 0.0058* | -0.0013 | 0.2466 |
| Inferior longitudinal fasciculus (left) | -0.0008 | 0.0645 | -0.0035 | 0.0037* | -0.0025 | 0.0244 |
| Inferior longitudinal fasciculus (right) | -0.0010 | 0.0071* | -0.0039 | 0.0007* | -0.0018 | 0.0871 |
| Middle cerebellar peduncle | 0.0003 | 0.5812 | 0.0001 | 0.9408 | -0.0005 | 0.7385 |
| Medial lemniscus (left) | 0.0004 | 0.4126 | 0.0001 | 0.9293 | -0.0006 | 0.6530 |
| Medial lemniscus (right) | -0.0007 | 0.1451 | -0.0025 | 0.0945 | -0.0024 | 0.0870 |
| Posterior thalamic radiation (left) | -0.0015 | 0.0005* | -0.0042 | 0.0007* | -0.0024 | 0.0405 |
| Posterior thalamic radiation (right) | -0.0015 | 0.0002* | -0.0041 | 0.0006* | -0.0021 | 0.0717 |
| Superior longitudinal fasciculus (left) | -0.0003 | 0.4409 | -0.0021 | 0.0906 | -0.0013 | 0.2874 |
| Superior longitudinal fasciculus (right) | -0.0005 | 0.1967 | -0.0011 | 0.3841 | 0.0000 | 0.9968 |
| Superior thalamic radiation (left) | -0.0008 | 0.0260 | -0.0005 | 0.6601 | 0.0000 | 0.9938 |
| Superior thalamic radiation (right) | -0.0010 | 0.0086* | -0.0003 | 0.7512* | 0.0006 | 0.5513 |
| Uncinate fasciculus (left) | -0.0009 | 0.0831 | 0.0008 | 0.5909 | 0.0012 | 0.3756 |
| Uncinate fasciculus (right) | -0.0012 | 0.0045* | -0.0019 | 0.1284 | -0.0001 | 0.9613 |

*Note.* $b_1/p_1$: Unstandardized Betas and *p* values for depression status derived from multiple regression analyses in the comparison of healthy and depressed participants without current symptoms; $b_1/p_1$: Unstandardized Betas and *p* values for depression status derived from multiple regression analyses in the comparison of healthy and depressed participants with current symptoms; $b_1/p_1$: Unstandardized Betas and *p* values for depression status derived from multiple regression analyses in the comparison of depressed participants with and without current symptoms. *Significant after correction for multiple comparisons (Benjamini-Hochberg procedure, false discovery rate: 5 %)

### 2.4 Sample overlap analyses

**Table T15** Partial correlations between polygenic risk for depression and simulated variables (depressed and healthy participants, Wray et al. (2018))

| | 1.00 | 0.5 | 0.2 | 0.1 | 0.05 | 0.01 | 1E-3 | 1E-4 | 1E-6 | 5E-8 |
|---|---|---|---|---|---|---|---|---|---|---|
| **0.1** | .0169 | .017 | .0171 | .0167 | .016 | .0135 | .0094 | .0067 | .0036 | .0034 |
| **0.2** | .0335 | .0336 | .0338 | .0331 | .0319 | .027 | .0191 | .0135 | .007 | .0065 |
| **0.3** | .0503 | .0504 | .0506 | .0497 | .0478 | .0405 | .0288 | .0203 | .0104 | .0096 |
| **0.4** | .0672 | .0673 | .0675 | .0664 | .0639 | .0542 | .0385 | .0272 | .0138 | .0127 |
| **0.5** | .0843 | .0845 | .0846 | .0833 | .0802 | .068 | .0485 | .0342 | .0173 | .0159 |
| **0.6** | .1017 | .1018 | .102 | .1004 | .0968 | .082 | .0585 | .0412 | .0208 | .0191 |
| **0.7** | .1194 | .1196 | .1197 | .1179 | .1136 | .0964 | .0688 | .0485 | .0244 | .0224 |
| **0.8** | .1375 | .1377 | .1378 | .1358 | .1309 | .111 | .0793 | .0559 | .028 | .0258 |
| **0.9** | .1561 | .1563 | .1564 | .1542 | .1486 | .1261 | .0901 | .0635 | .0318 | .0292 |

**Table T16** Partial correlations between polygenic risk for depression and simulated variables (healthy participants only, Wray et al. (2018))

| | 1.00 | 0.5 | 0.2 | 0.1 | 0.05 | 0.01 | 1E-3 | 1E-4 | 1E-6 | 5E-8 |
|---|---|---|---|---|---|---|---|---|---|---|
| **0.1** | < -.0001 | < -.0001 | < -.0001 | < -.0001 | < -.0001 | -.0001 | -.0002 | -.0002 | < -.0001 | < -.0001 |
| **0.2** | < -.0001 | < -.0001 | < -.0001 | < -.0001 | < -.0001 | -.0001 | -.0002 | -.0002 | < -.0001 | < -.0001 |
| **0.3** | < -.0001 | < -.0001 | < -.0001 | < -.0001 | < -.0001 | -.0001 | -.0002 | -.0002 | < -.0001 | < -.0001 |
| **0.4** | < -.0001 | < -.0001 | < -.0001 | < -.0001 | < -.0001 | -.0001 | -.0002 | -.0002 | < -.0001 | < -.0001 |
| **0.5** | < -.0001 | < -.0001 | < -.0001 | < -.0001 | < -.0001 | -.0001 | -.0002 | -.0002 | < -.0001 | < -.0001 |
| **0.6** | < -.0001 | < -.0001 | < -.0001 | < -.0001 | < -.0001 | -.0001 | -.0002 | -.0002 | < -.0001 | < -.0001 |
| **0.7** | < -.0001 | < -.0001 | < -.0001 | < -.0001 | < -.0001 | -.0001 | -.0002 | -.0002 | < -.0001 | < -.0001 |
| **0.8** | < -.0001 | < -.0001 | < -.0001 | < -.0001 | < -.0001 | -.0001 | -.0002 | -.0002 | < -.0001 | < -.0001 |
| **0.9** | < -.0001 | < -.0001 | < -.0001 | < -.0001 | < -.0001 | -.0001 | -.0002 | -.0002 | < -.0001 | < -.0001 |

**Table T17** Partial correlations between polygenic risk for depression and simulated variables (depressed and healthy participants, Howard et al. (2019))

| | 1.00 | 0.5 | 0.2 | 0.1 | 0.05 | 0.01 | 1E-3 | 1E-4 | 1E-6 | 5E-8 |
|---|---|---|---|---|---|---|---|---|---|---|
| **0.1** | .0349 | .0348 | .0339 | .0326 | .0309 | .0265 | .0197 | .0129 | .007 | .0048 |
| **0.2** | .0696 | .0694 | .0675 | .065 | .0617 | .0529 | .0392 | .026 | .014 | .0093 |
| **0.3** | .1045 | .1042 | .1014 | .0976 | .0927 | .0793 | .0588 | .0391 | .021 | .0137 |
| **0.4** | .1398 | .1394 | .1356 | .1305 | .124 | .1061 | .0785 | .0524 | .0281 | .0183 |
| **0.5** | .1755 | .175 | .1702 | .1639 | .1557 | .1331 | .0985 | .0658 | .0352 | .0228 |
| **0.6** | .2117 | .2111 | .2054 | .1977 | .1879 | .1606 | .1189 | .0795 | .0425 | .0275 |
| **0.7** | .2487 | .248 | .2412 | .2322 | .2207 | .1886 | .1396 | .0934 | .0499 | .0322 |
| **0.8** | .2864 | .2857 | .2778 | .2675 | .2543 | .2173 | .1608 | .1076 | .0575 | .037 |
| **0.9** | .3252 | .3243 | .3155 | .3037 | .2887 | .2466 | .1825 | .1222 | .0652 | .042 |

**Table T18** Partial correlations between polygenic risk for depression and simulated variables (healthy participants only, Howard et al. (2019))

| | **1.00** | **0.5** | **0.2** | **0.1** | **0.05** | **0.01** | **1E-3** | **1E-4** | **1E-6** | **5E-8** |
|---|---|---|---|---|---|---|---|---|---|---|
| **0.1** | .0001 | .0001 | .0001 | < .0001 | < -.0001 | < -.0001 | -.0001 | -.0002 | < -.0001 | .0001 |
| **0.2** | .0001 | .0001 | .0001 | < .0001 | < -.0001 | < -.0001 | -.0001 | -.0002 | < -.0001 | .0001 |
| **0.3** | .0001 | .0001 | .0001 | < .0001 | < -.0001 | < -.0001 | -.0001 | -.0002 | < -.0001 | .0001 |
| **0.4** | .0001 | .0001 | .0001 | < .0001 | < -.0001 | < -.0001 | -.0001 | -.0002 | < -.0001 | .0001 |
| **0.5** | .0001 | .0001 | .0001 | < .0001 | < -.0001 | < -.0001 | -.0001 | -.0002 | < -.0001 | .0001 |
| **0.6** | .0001 | .0001 | .0001 | < .0001 | < -.0001 | < -.0001 | -.0001 | -.0002 | < -.0001 | .0001 |
| **0.7** | .0001 | .0001 | .0001 | < .0001 | < -.0001 | < -.0001 | -.0001 | -.0002 | < -.0001 | .0001 |
| **0.8** | .0001 | .0001 | .0001 | < .0001 | < -.0001 | < -.0001 | -.0001 | -.0002 | < -.0001 | .0001 |
| **0.9** | .0001 | .0001 | .0001 | < .0001 | < -.0001 | < -.0001 | -.0001 | -.0002 | < -.0001 | .0001 |

**Table T19** Partial correlations between polygenic risk for depression and simulated variables (depressed and healthy participants, Baselmans et al. (2019))

| | **1.00** | **0.5** | **0.2** | **0.1** | **0.05** | **0.01** | **1E-3** | **1E-4** | **1E-6** | **5E-8** |
|---|---|---|---|---|---|---|---|---|---|---|
| **0.1** | .0268 | .0267 | .026 | .0251 | .0238 | .0206 | .0153 | .0113 | .0076 | .0057 |
| **0.2** | .0535 | .0534 | .0519 | .0503 | .0475 | .041 | .0306 | .0226 | .0151 | .0113 |
| **0.3** | .0805 | .0802 | .078 | .0756 | .0714 | .0616 | .046 | .034 | .0227 | .017 |
| **0.4** | .1077 | .1073 | .1043 | .1011 | .0955 | .0824 | .0615 | .0455 | .0304 | .0227 |
| **0.5** | .1352 | .1347 | .131 | .1269 | .1199 | .1034 | .0772 | .0571 | .0381 | .0285 |
| **0.6** | .1631 | .1626 | .1581 | .1532 | .1447 | .1247 | .0932 | .0689 | .046 | .0343 |
| **0.7** | .1916 | .191 | .1857 | .1799 | .1699 | .1465 | .1094 | .0809 | .054 | .0403 |
| **0.8** | .2208 | .2201 | .2139 | .2073 | .1957 | .1687 | .1261 | .0932 | .0622 | .0464 |
| **0.9** | .2507 | .2499 | .2429 | .2354 | .2222 | .1916 | .1432 | .1058 | .0706 | .0527 |

**Table T20** Partial correlations between polygenic risk for depression and simulated variables (healthy participants only, Baselmans et al. (2019))

| | **1.00** | **0.5** | **0.2** | **0.1** | **0.05** | **0.01** | **1E-3** | **1E-4** | **1E-6** | **5E-8** |
|---|---|---|---|---|---|---|---|---|---|---|
| **0.1** | < -.0001 | < -.0001 | < -.0001 | < -.0001 | < -.0001 | -.0001 | -.0002 | -.0001 | -.0001 | -.0001 |
| **0.2** | < -.0001 | < -.0001 | < -.0001 | < -.0001 | < -.0001 | -.0001 | -.0002 | -.0001 | -.0001 | -.0001 |
| **0.3** | < -.0001 | < -.0001 | < -.0001 | < -.0001 | < -.0001 | -.0001 | -.0002 | -.0001 | -.0001 | -.0001 |
| **0.4** | < -.0001 | < -.0001 | < -.0001 | < -.0001 | < -.0001 | -.0001 | -.0002 | -.0001 | -.0001 | -.0001 |
| **0.5** | < -.0001 | < -.0001 | < -.0001 | < -.0001 | < -.0001 | -.0001 | -.0002 | -.0001 | -.0001 | -.0001 |
| **0.6** | < -.0001 | < -.0001 | < -.0001 | < -.0001 | < -.0001 | -.0001 | -.0002 | -.0001 | -.0001 | -.0001 |
| **0.7** | < -.0001 | < -.0001 | < -.0001 | < -.0001 | < -.0001 | -.0001 | -.0002 | -.0001 | -.0001 | -.0001 |
| **0.8** | < -.0001 | < -.0001 | < -.0001 | < -.0001 | < -.0001 | -.0001 | -.0002 | -.0001 | -.0001 | -.0001 |
| **0.9** | < -.0001 | < -.0001 | < -.0001 | < -.0001 | < -.0001 | -.0001 | -.0002 | -.0001 | -.0001 | -.0001 |

*Note.* The tables 15 to 20 show the partial correlations for the different correlations strengths between simulated variables and depression (0.1 to 0.9) and for the different polygenic risk thresholds. The data presented here is also presented in main figure 4AB.

**Figure S2** Partial correlation between simulated variables (correlated with depression status) and polygenic risk scores

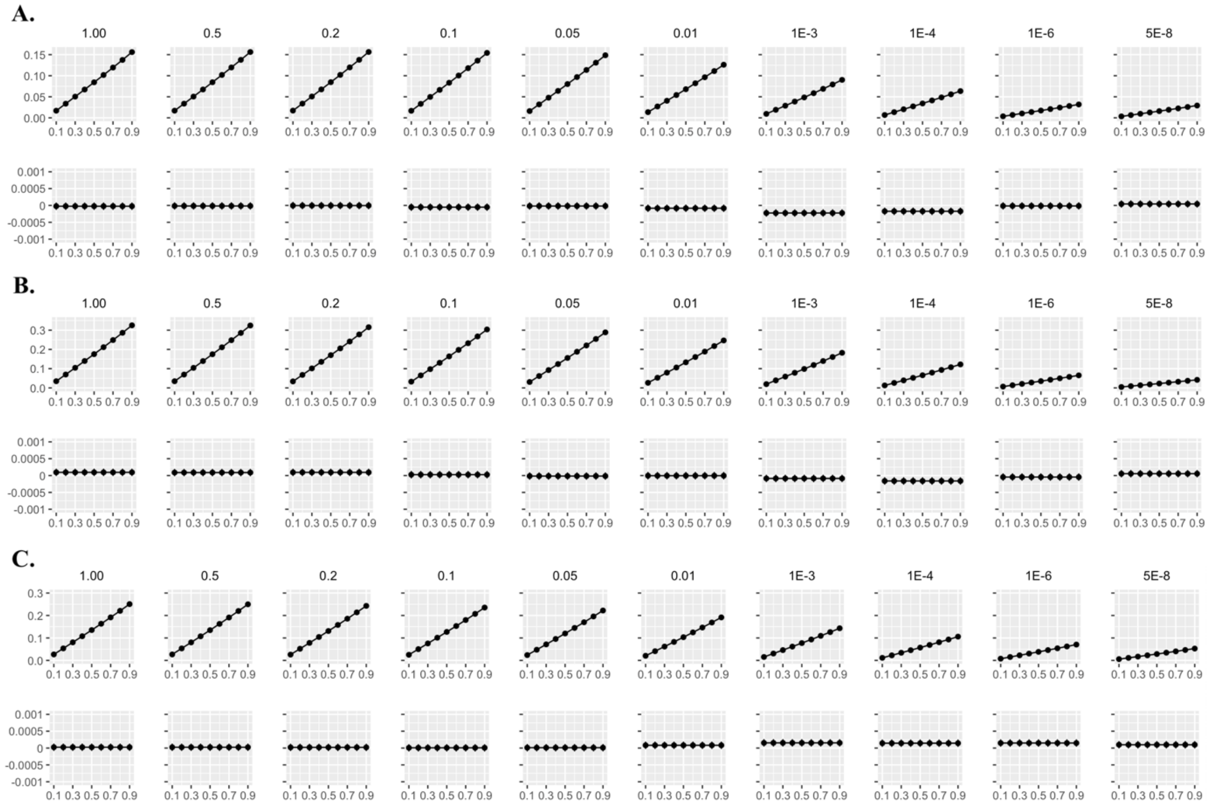


*Note*. Figure A shows the partial correlation for the polygenic risk scores based on Wray et al. (2018), B for the polygenic risk scores based on Howard et al. (2019) and C for the polygenic risk score based on Baselmans et al. (2019). Upper figures include healthy and depressed participants, lower figures only healthy participants. A more compact depiction is presented in main figure 4AB.

**Figure S3: Detailed results from the non-parametric test**

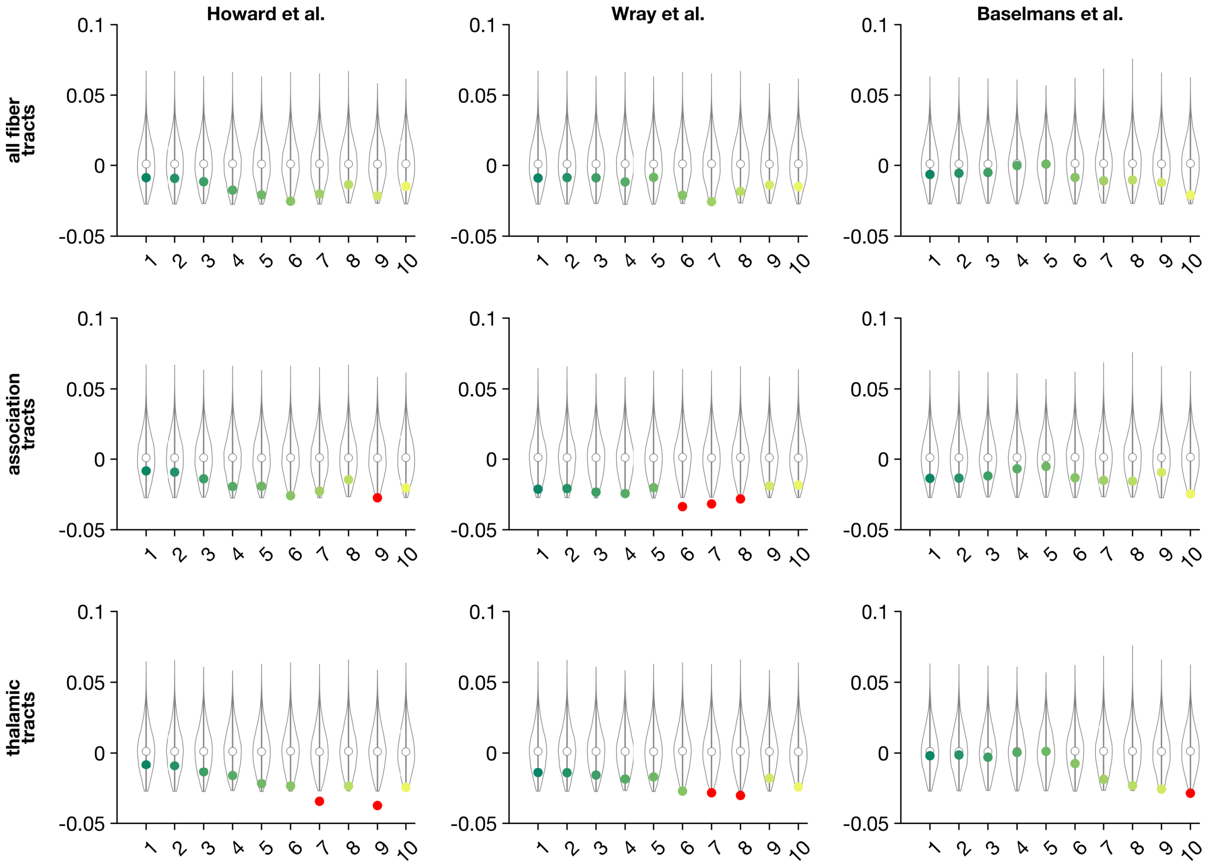


*Note: The violins summarize the distribution of random PGS associations (largest 9950, lower cutoff at p<.05) for each fiber tract group (in rows), each PGS (in columns), and each threshold (x-axis). Empirical PGS associations are superimposed in shades of green, and highlighted in red when significantly smaller (i.e. more substantial) than in comparable random data (p<.05).*

### 2.5 Genetic correlations

**Table T21** SNP-based heritabilities for FA phenotypes

| Tract type/ Fiber tract | $h^2_{SNP}$ | *SE* |
|---|---|---|
| PC derived from all fiber tract | 0.320 | 0.030 |
| PC derived from association/commissural fibers | 0.318 | 0.030 |
| PC derived from thalamic radiations | 0.296 | 0.027 |
| Anterior thalamic radiation (right) | 0.281 | 0.025 |
| Cingulate gyrus part of cingulum (left) | 0.191 | 0.021 |
| Forceps minor | 0.327 | 0.028 |
| Inferior fronto-occipital fasciculus (right) | 0.311 | 0.027 |
| Inferior longitudinal fasciculus (left) | 0.248 | 0.028 |
| Inferior longitudinal fasciculus (right) | 0.276 | 0.028 |
| Posterior thalamic radiation (left) | 0.212 | 0.025 |
| Posterior thalamic radiation (right) | 0.208 | 0.023 |
| Superior thalamic radiation (left) | 0.239 | 0.026 |
| Superior thalamic radiation (right) | 0.241 | 0.023 |
| Uncinate fasciculus (right) | 0.263 | 0.023 |

**Table T22** Genetic correlations between depression and FA of individual fiber tracts

| Tract type | GWAS Depression | *rg* | *se (rg)* | *z* | *p* |
|---|---|---|---|---|---|
| Anterior thalamic radiation (right) | Wray et al. (2018) | -.04 | .037 | -1.056 | .291 |
| | Howard et al. (2018) | -.027 | .034 | -0.801 | .422 |
| | Baselmans et al. (2019) | -.023 | .035 | -0.654 | .513 |
| Cingulate gyrus part of cingulum (left) | Wray et al. (2018) | -.118 | .043 | -2.774 | .006 |
| | Howard et al. (2018) | -.08 | .037 | -2.109 | .035 |
| | Baselmans et al. (2019) | -.033 | .047 | -0.67 | .484 |
| Forceps minor | Wray et al. (2018) | -.095 | .036 | -2.664 | .008 |
| | Howard et al. (2018) | -.058 | .032 | -1.809 | .07 |
| | Baselmans et al. (2019) | -.054 | .033 | -1.621 | .105 |
| Inferior fronto-occipital fasciculus (right) | Wray et al. (2018) | -.02 | .034 | -0.58 | .562 |
| | Howard et al. (2018) | -.029 | .03 | -0.946 | .344 |
| | Baselmans et al. (2019) | -.01 | .035 | 0.278 | .781 |
| Inferior longitudinal fasciculus (left) | Wray et al. (2018) | <.001 | .037 | 0.011 | .991 |
| | Howard et al. (2018) | .009 | .035 | 0.266 | .79 |
| | Baselmans et al. (2019) | 0.01 | .04 | 0.251 | .802 |
| Inferior longitudinal fasciculus (right) | Wray et al. (2018) | -.028 | .036 | -0.758 | .448 |
| | Howard et al. (2018) | .006 | .033 | 0.193 | .847 |
| | Baselmans et al. (2019) | .013 | .037 | -0.254 | .724 |
| Posterior thalamic radiation (left) | Wray et al. (2018) | -.048 | .039 | -1.209 | .227 |
| | Howard et al. (2018) | -.066 | .034 | -1.943 | .052 |
| | Baselmans et al. (2019) | -.075 | .038 | -1.991 | .047 |
| Posterior thalamic radiation (right) | Wray et al. (2018) | -.018 | .042 | -0.432 | .666 |
| | Howard et al. (2018) | -.037 | .032 | -1.136 | .256 |
| | Baselmans et al. (2019) | -.037 | .038 | -0.975 | .33 |
| Superior thalamic radiation (left) | Wray et al. (2018) | -.046 | .04 | -1.172 | .241 |
| | Howard et al. (2018) | .007 | .034 | 0.197 | .844 |
| | Baselmans et al. (2019) | .013 | .039 | 0.337 | .736 |
| Superior thalamic radiation (right) | Wray et al. (2018) | -.052 | .039 | -1.34 | .181 |
| | Howard et al. (2018) | -.019 | .034 | -0.553 | .58 |
| | Baselmans et al. (2019) | -.007 | .039 | 0.172 | .863 |
| Uncinate fasciculus (right) | Wray et al. (2018) | -.037 | .036 | -1.032 | .302 |
| | Howard et al. (2018) | -.05 | .03 | -1.637 | .102 |
| | Baselmans et al. (2019) | -.022 | .037 | 0.593 | .553 |
